# Vaginal metabolome signatures of high-risk HPV infection trajectories in HIV-negative premenopausal women

**DOI:** 10.64898/2026.04.21.26351401

**Authors:** Clement A. Adebamowo, Sally Nneoma Adebamowo, Gbolahan Toluwalope, Ogechukwu Ikwueme, Ayo Famooto, Yinka Owoade, ACCME Research Group as part of the H3Africa Consortium

## Abstract

Persistent detection of high-risk human papillomavirus (HPV) is required for cervical carcinogenesis, yet the metabolic phenotype associated with distinct HPV transition states remains incompletely defined. We analyzed vaginal metabolomics data from 71 HIV-negative, non-smoking, premenopausal women without other sexually transmitted infections, grouped by three-visit HPV trajectories: persistent negative (NNN, n=20), late incident positivity (NNP, n=9), conversion with persistence (NPP, n=13), clearance after prior positivity (PPN, n=16), and persistent positive (PPP, n=13). After detection-based filtering, 186 putative and 64 quantitatively estimated metabolites were retained for integrated univariate, multivariate, network, pathway, and machine learning analyses. Global class separation was weak by PERMANOVA and by five-class classification, indicating that the vaginal metabolome does not reorganize broadly across all HPV states. In contrast, trajectory-specific signals were reproducible. The strongest pairwise contrast was NNP versus PPP (best cross-validated ROC AUC 0.778; permutation p=0.039). Glycolic acid was the dominant single metabolite, particularly for NNP versus PPP (Mann–Whitney p=6.96×10^-4, FDR=0.0446, AUROC=0.902; detection 88.9% versus 15.4%; combined abundance+detection FDR=0.0010). Persistent positivity was characterized by a focused uracil-high, methyl-donor/redox-low signature, including lower glycolic acid, S-adenosylmethionine, NAD+, and betaine, together with higher uracil. Ratio mining further sharpened discrimination, with uracil/S-adenosylmethionine and uracil/creatinine among the best PPP classifiers, and glucose 1-phosphate/isovaleric acid-valeric acid strongly separating NNP from NPP. These data support a model in which HPV trajectory is encoded by targeted metabolic states rather than a diffuse HPV-positive versus HPV-negative metabolomic shift.

## Introduction

High-risk HPV is the necessary cause of nearly all cervical cancers, but only a subset of infections persist and progress ^1-3^. The biological interval between viral acquisition, persistence, clearance, and neoplastic progression is therefore the most informative window for biomarker discovery. Most studies have emphasized virology, host immunity, or the vaginal microbiota, while relatively fewer have interrogated the vaginal metabolome as an integrated readout of epithelial physiology, host-microbial interactions, substrate availability, and inflammatory tone.

The reproductive-age vaginal ecosystem is dynamic and highly structured, with strong links between microbial community state, glycogen processing, lactic acid production, pH, and mucosal immunity ^4-14^. Longitudinal microbiome studies further suggest that HPV detection is not simply a binary event, but a process embedded within temporally changing cervicovaginal states ^9,15,16^. Systematic reviews and network meta-analyses support associations between non-optimal vaginal community structure, HPV, and cervical dysplasia, although directionality and mechanism remain unresolved ^6,7,16-19^.

Metabolomics is appealing in this context because it can capture both stable pathway-level features and subtle transition-state signals. In bacterial vaginosis and other genital tract conditions, vaginal metabolites strongly reflect microbial composition and host chemistry, including lactic acid, amino acid catabolites, biogenic amines, and redox-linked intermediates ^13,20^. Whether a comparable, yet more nuanced, signal tracks high-risk HPV acquisition, persistence, or clearance in an otherwise tightly phenotyped healthy population has been less clear.

Here, we analyze a vaginal metabolomics dataset from HIV-negative, premenopausal, non-smoking women free of other sexually transmitted infections. The cohort was classified into five longitudinal HPV trajectories spanning persistent negativity, late incident positivity, conversion with persistence, clearance after repeated positivity, and persistent positivity (Figure 1). The central question was not only whether HPV-positive women differ from HPV-negative women, but whether distinct trajectory classes encode interpretable metabolic states. We therefore prioritized multi-level inference: individual metabolites, pairwise class contrasts, grouped comparisons, metabolite ratios, module and pathway structure, and machine-learning-based discrimination in this study.

**Fig. 1.**
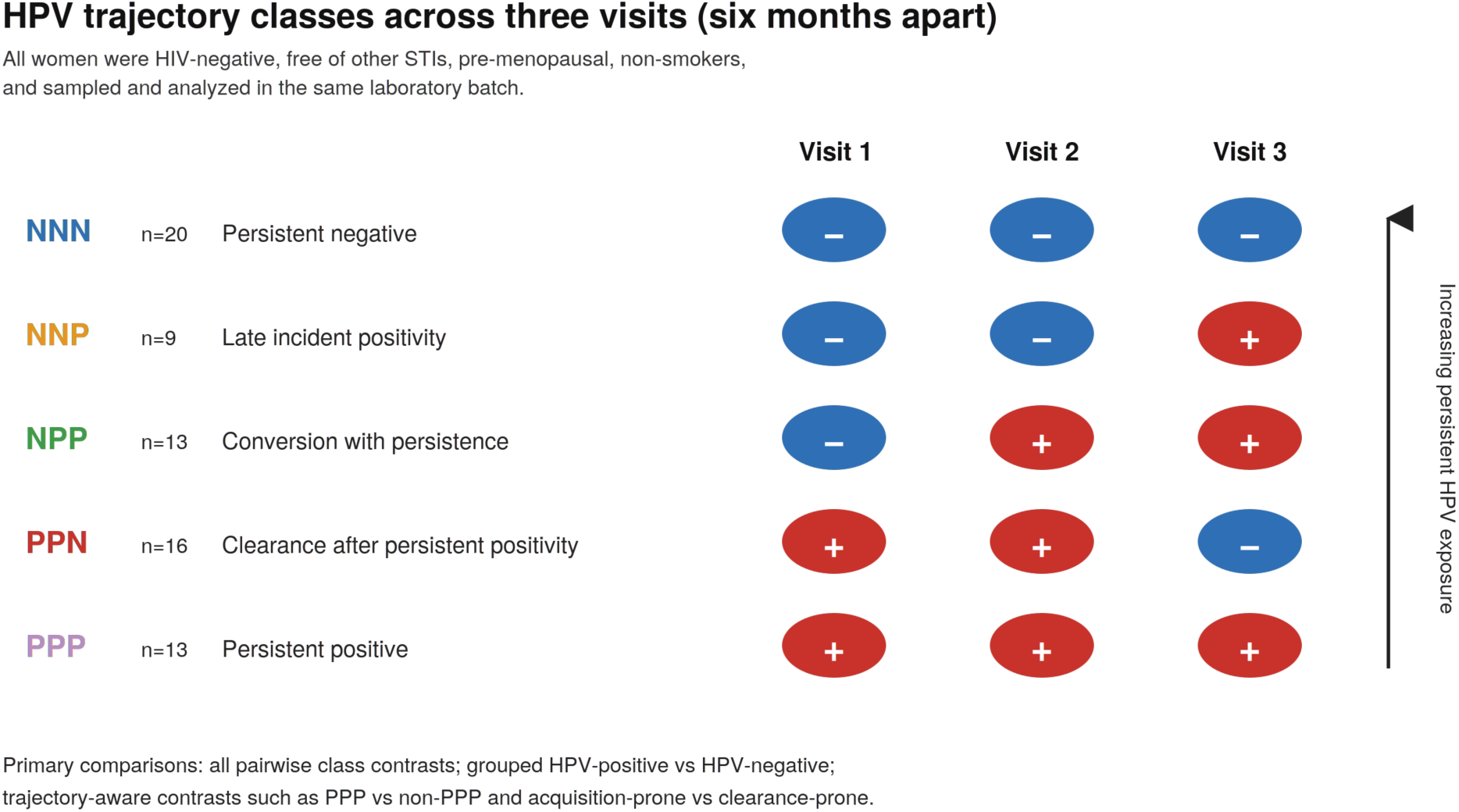
Study design and HPV trajectory class definitions. Women were grouped according to three serial high-risk HPV measurements obtained six months apart.

## Results

### Cohort composition and analytic overview

The study included 71 women divided into five trajectory classes: NNN (n=20), NNP (n=9), NPP (n=13), PPN (n=16), and PPP (n=13) (Figure 1). The mean age of the cohort was 37 years, all participants were HIV negative, free of sexually transmitted infections (STI), premenopausal, non-smoking, sampled contemporaneously, and analytically processed together. There were 254 putative and 116 quantitatively estimated metabolites and in the validated downstream analysis, we retained 186 putative and 64 quantified metabolites that had adequate detection for robust statistical modeling (Table 1). The detection burden differed modestly across classes, but these differences did not translate into broad class-level metabolome separation (Supplementary Figures. 1 and 2).

**Table 1.** Cohort and analytic overview.

| Metric | Value |
| --- | --- |
| Participants | 71 |
| Trajectory classes | NNN=20; NNP=9; NPP=13; PPN=16; PPP=13 |
| Putative metabolites retained | 186 / 254 |
| Quantified metabolites retained | 64 / 116 |
| PERMANOVA putative | $R^2=0.054$, $p=0.548$ |
| PERMANOVA quantitative | $R^2=0.043$, $p=0.790$ |
| Best 5-class model | LinearSVM (Combined); balanced accuracy=0.235 |
| Best pairwise contrast | NNP vs PPP; AUC=0.778 |
| Best grouped binary contrast | PPP vs non-PPP; AUC=0.646 |

### Global metabolome structure is weakly separated across the five HPV classes

Principal component analysis (PCA) of both the putative and quantitative matrices showed substantial overlap among classes, consistent with the weak between-group structure observed by PERMANOVA (putative R²=0.0536, p=0.548; quantitative R²=0.0433, p=0.790) (Table 1, Supplementary Figures 3 and 4). Likewise, multiclass machine learning was only modestly informative. The best five-class model achieved a balanced accuracy of 0.235 using a combined feature set and a linear support vector machine (Figure 2). Thus, the primary signal in this cohort is not a globally reorganized five-state metabolome.

**Figure 2:**
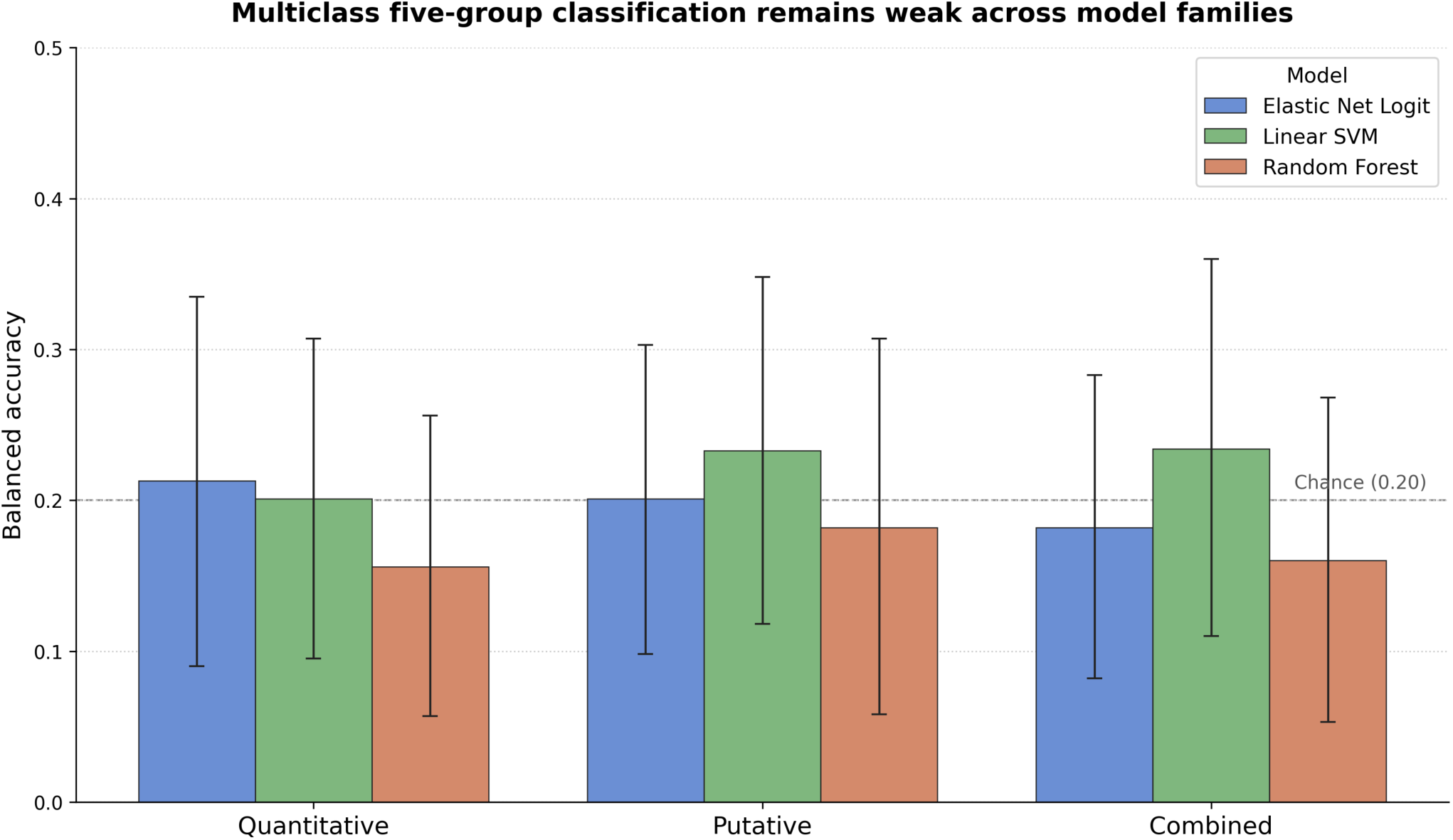
Five-class classification remained weak across model families and feature layers.

Even within this largely overlapping global structure, geometry in latent space was not random. Quantitative centroid distances were largest between NNP and PPP (3.73), whereas NNN and PPP were comparatively close (1.93), suggesting that persistent positivity is not simply the metabolomic opposite of persistent negativity. Instead, the most distinctive state appeared to be late incident positivity (NNP), which repeatedly occupied the most distant position from other class centroids (Supplementary Figures 5 and 6). Notably, several metabolites with high unsupervised PCA loadings, including lactic acid and low-abundance SCFA/keto-acid features, did not translate into reproducible HPV trajectory markers, indicating that the dominant latent variance in CF71 is largely orthogonal to HPV-associated variation.

### Trajectory-specific pairwise contrasts outperform grouped HPV-positive versus HPV-negative comparisons

The best-performing pairwise comparison was NNP versus PPP, with a cross-validated ROC AUC of 0.778 and balanced accuracy of 0.683 using elastic-net logistic regression on combined features (Table 2, Figure 3). The next strongest contrast was NNP versus NPP (AUC 0.722), indicating that the metabolic distinction of NNP is retained even relative to another transition class. By contrast, the best grouped binary comparison was PPP versus non-PPP (AUC 0.646), whereas “ever HPV positive versus NNN” and “final positive versus final negative” were weak. This pattern strongly suggests that longitudinal trajectory is more informative than collapsing women into broad HPV-positive or HPV-negative categories.

**Fig. 3.**
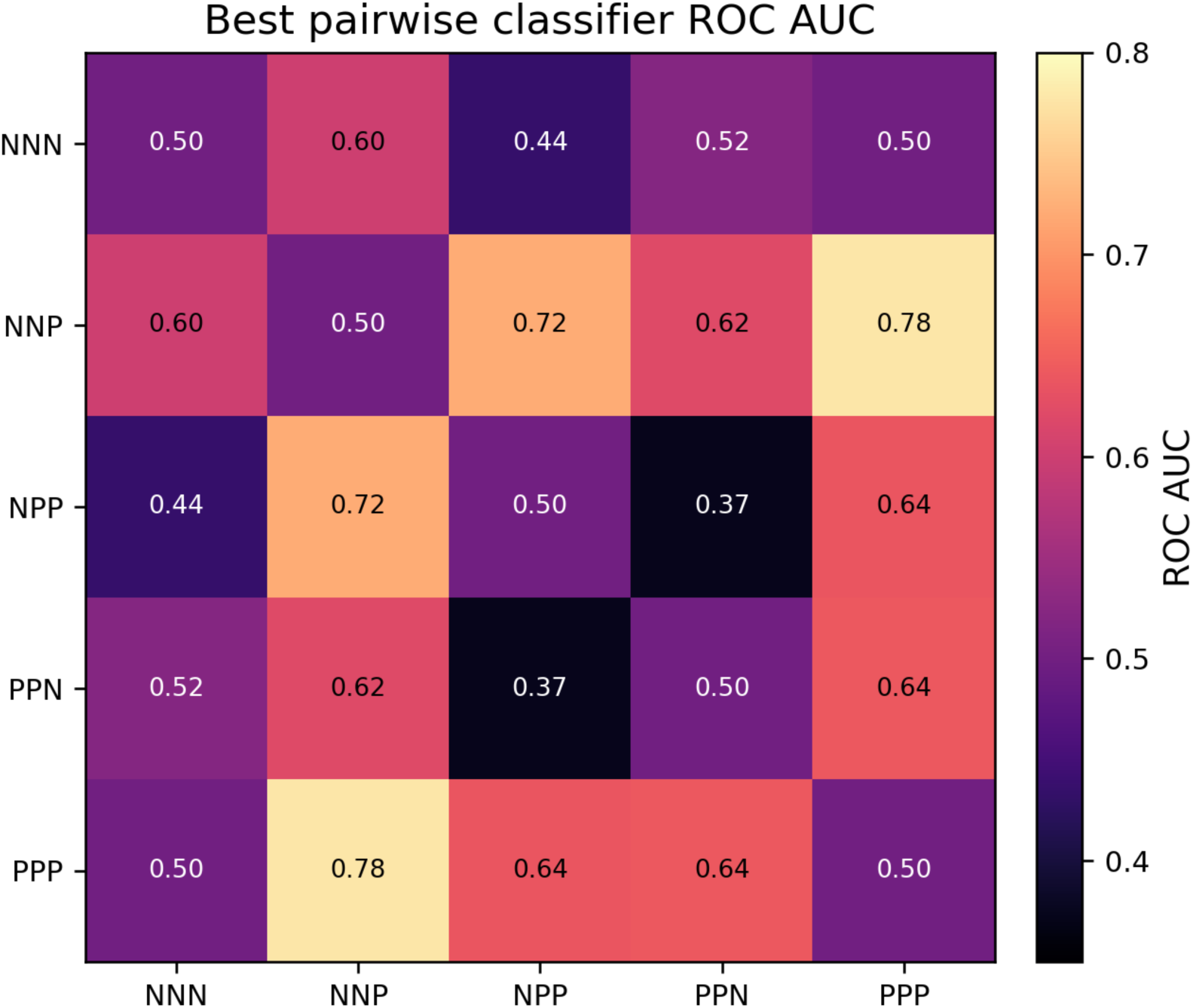
Heatmap of the best pairwise classifier ROC AUC values. The strongest contrast is NNP versus PPP.

**Table 2.** Top pairwise classifiers.

| Group 1 | Group 2 | Contrast | Layer | Model | Balanced accuracy | ROC AUC |
| --- | --- | --- | --- | --- | --- | --- |
| NNP | PPP | NNP vs PPP | Combined | ElasticNetLogit | 0.683 | 0.778 |
| NNP | NPP | NNP vs NPP | Putative | LinearSVM | 0.656 | 0.722 |
| PPN | PPP | PPN vs PPP | Putative | LinearSVM | 0.556 | 0.641 |
| NPP | PPP | NPP vs PPP | Combined | LinearSVM | 0.572 | 0.637 |
| NNP | PPN | NNP vs PPN | Putative | ElasticNetLogit | 0.608 | 0.622 |

Permutation analysis reinforced this interpretation. For the putative-layer NNP versus PPP classifier, the observed AUC exceeded the null distribution with an empirical p value of approximately 0.039, while PPP versus non-PPP did not show comparable permutation support (Figure 4). One-versus-rest analysis likewise indicated that NNP was the easiest class to detect (AUC 0.666), again emphasizing the biological distinctiveness of this late incident positivity state (Figure 5).

**Figure 4:**
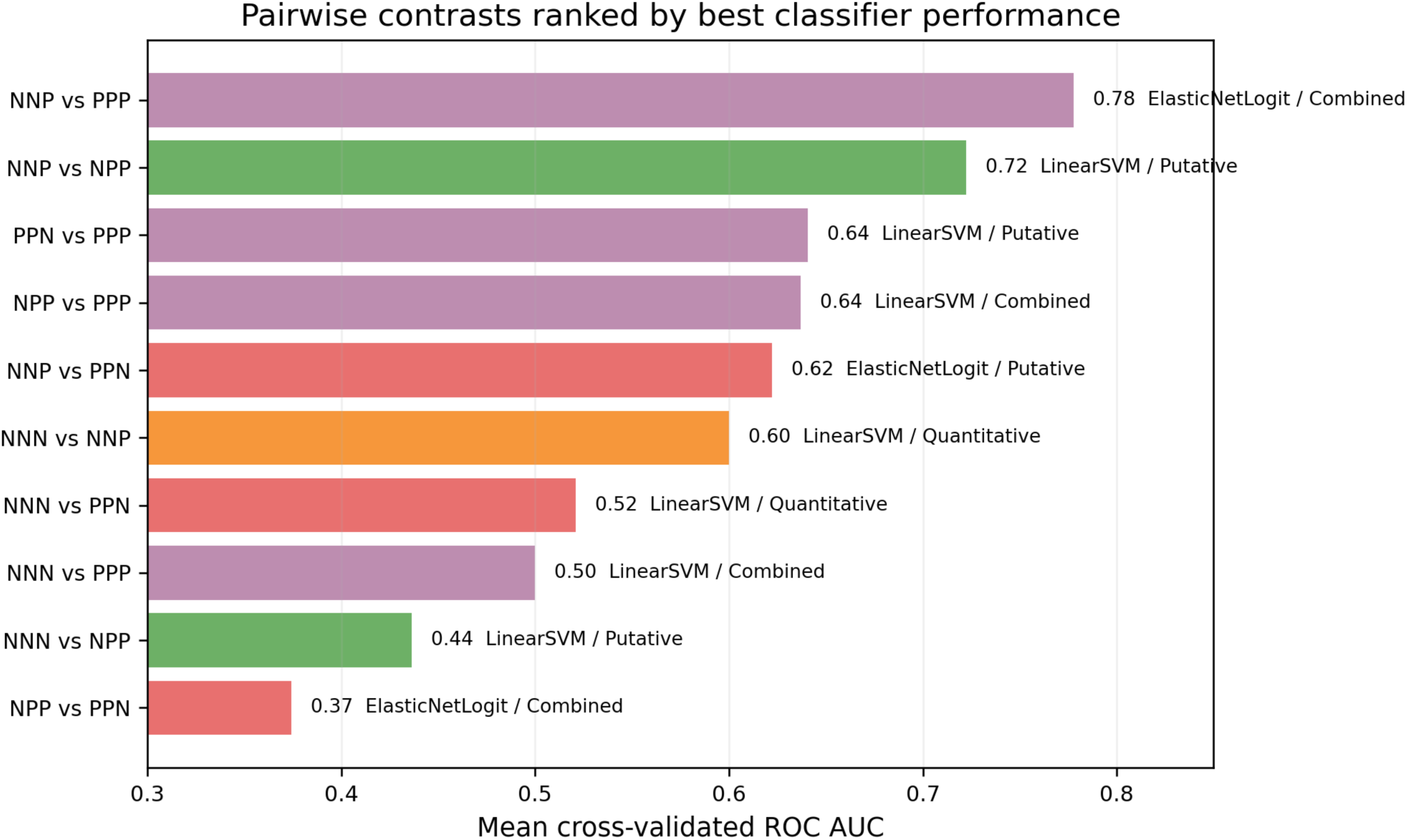
Pairwise contrast analysis by best classifier performance.

**Figure 5:**
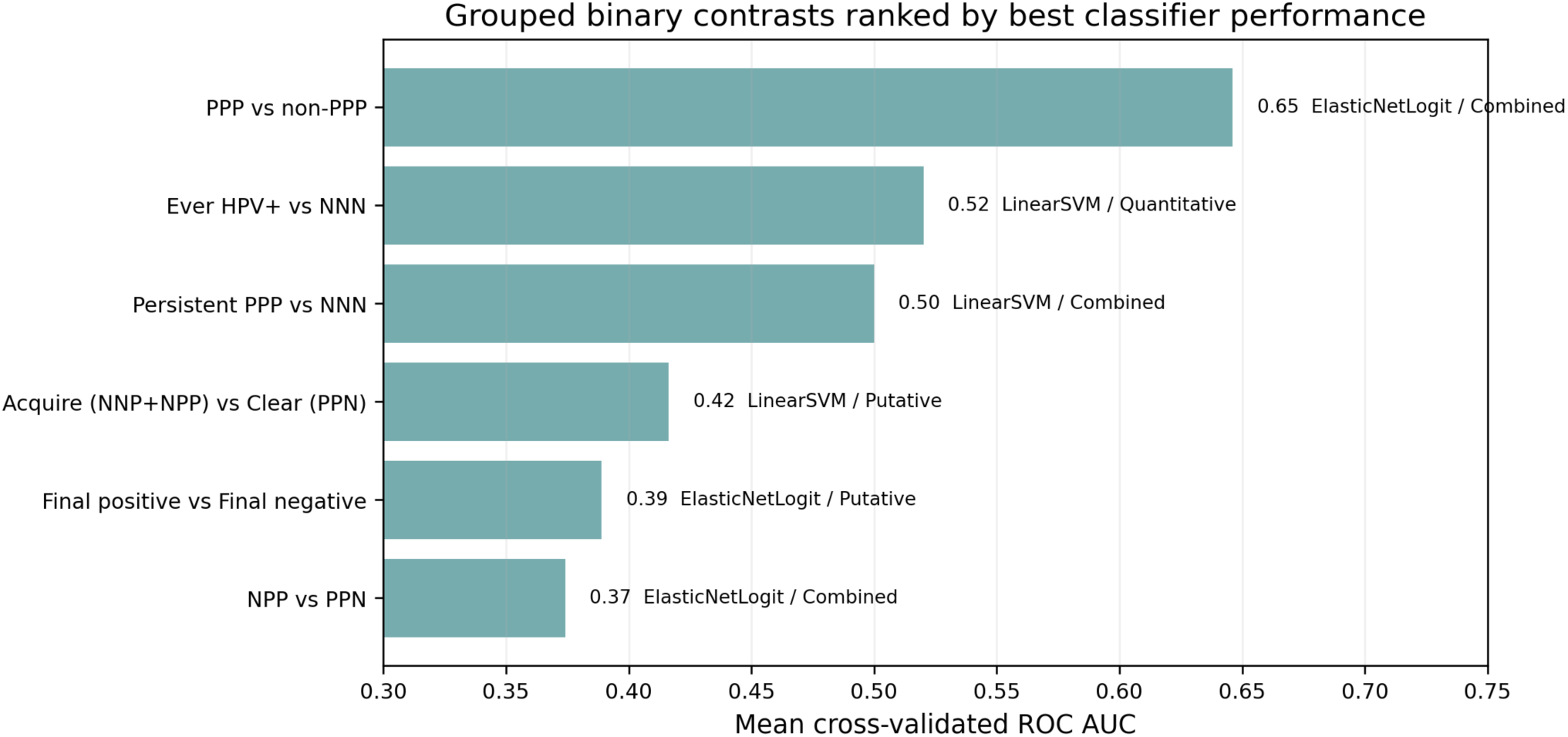
Grouped binary contrasts ranked by best classifier performance.

### Glycolic acid dominates the strongest pairwise signal, while persistent positivity is marked by a focused uracil-high and methyl-donor/redox-low state

At the single-metabolite level, glycolic acid was the clearest discriminator. In the strongest contrast (NNP versus PPP), glycolic acid showed a Mann–Whitney *p*-value of 6.96×10^-4^, an FDR of 0.0446, and an AUROC of 0.902, with substantially lower abundance in PPP than NNP (Figure 6, Table 3). The detection signal ran in the same direction. Glycolic acid was detected in 88.9% of NNP samples but only 15.4% of PPP samples (Fisher’s test *p*-value = 0.00155). When abundance and detection were integrated into a two-part evidence framework, glycolic acid remained the most robust marker (combined *p*-value = 1.6×10^-5^; combined FDR=0.0010).

**Figure 6:**
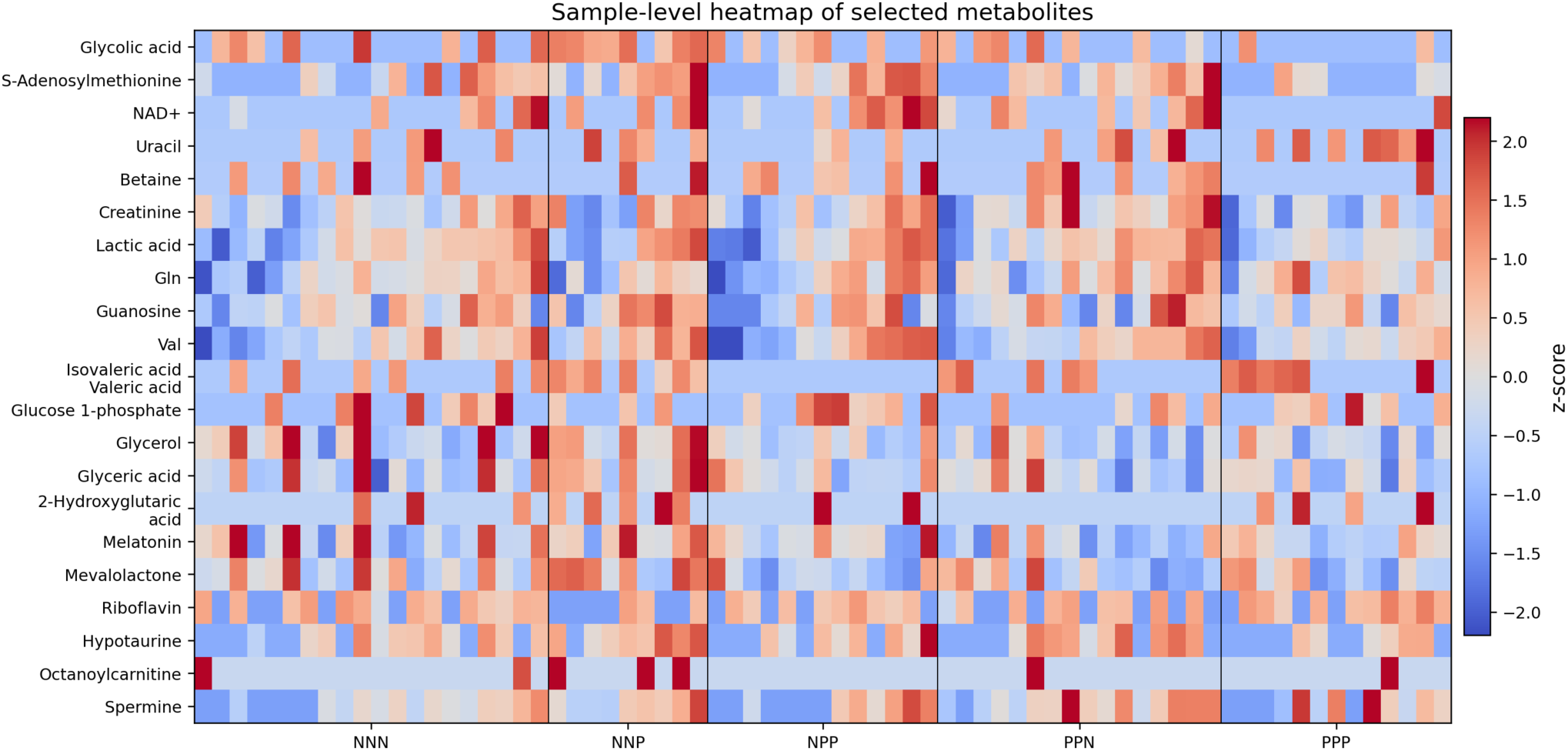
Single metabolite level heatmap of selected metabolites.

**Table 3.** Top quantitative markers for NNP versus PPP.

| Metabolite | Abundance<br>p | AUC | Detection p | Combined<br>FDR |
| --- | --- | --- | --- | --- |
| Glycolic acid | 0.000696 | 0.902 | 0.00155 | 0.00102 |
| S-Adenosylmethionine | 0.0116 | 0.812 | 0.0991 | 0.285 |
| 3-Hydroxybutyric acid | 0.0899 | 0.684 | 0.0743 | 0.745 |
| NAD<sup>+</sup> | 0.069 | 0.675 | 0.116 | 0.745 |
| Cytosine | 0.104 | 0.684 | 0.187 | 1 |

Beyond glycolic acid, the PPP state was characterized by a compact set of coordinated changes (Figure 7). Compared with NNP and, to a lesser extent, with non-PPP samples overall, PPP showed lower S-adenosylmethionine, lower NAD+, lower betaine, and higher uracil (Figures 8 and 9). The pattern suggests not a diffuse reprogramming of the vaginal metabolome, but a relatively low-dimensional state involving nucleotide handling together with attenuated methyl-donor and redox-associated metabolites. Quantitative group means further indicated that lactic acid and creatinine were comparatively stable across classes, underscoring the selectivity of the identified signal.

**Fig. 7.**
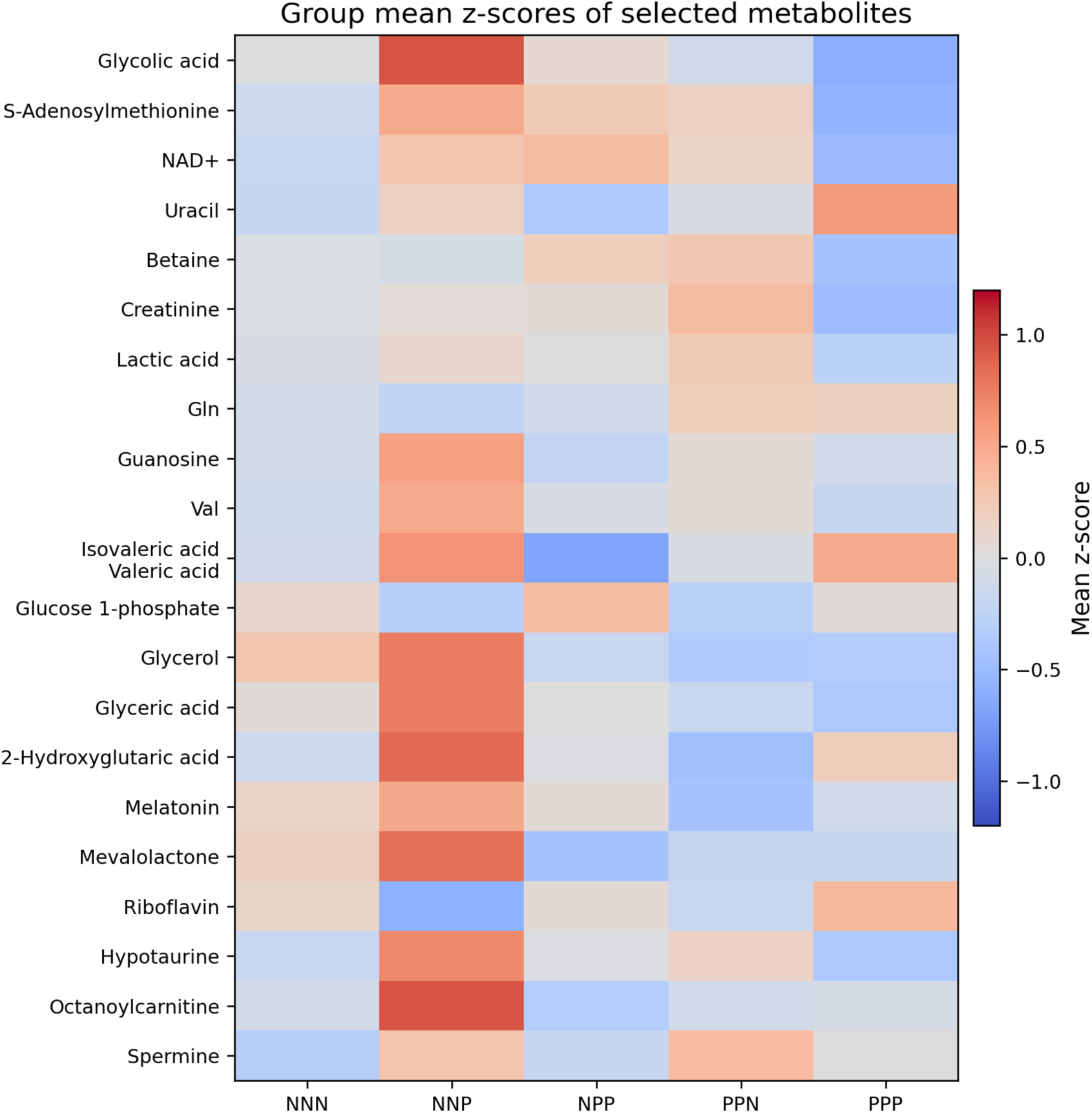
Group mean heatmap for selected metabolites highlights the focused PPP state and the distinctiveness of NNP.

**Fig. 8.**
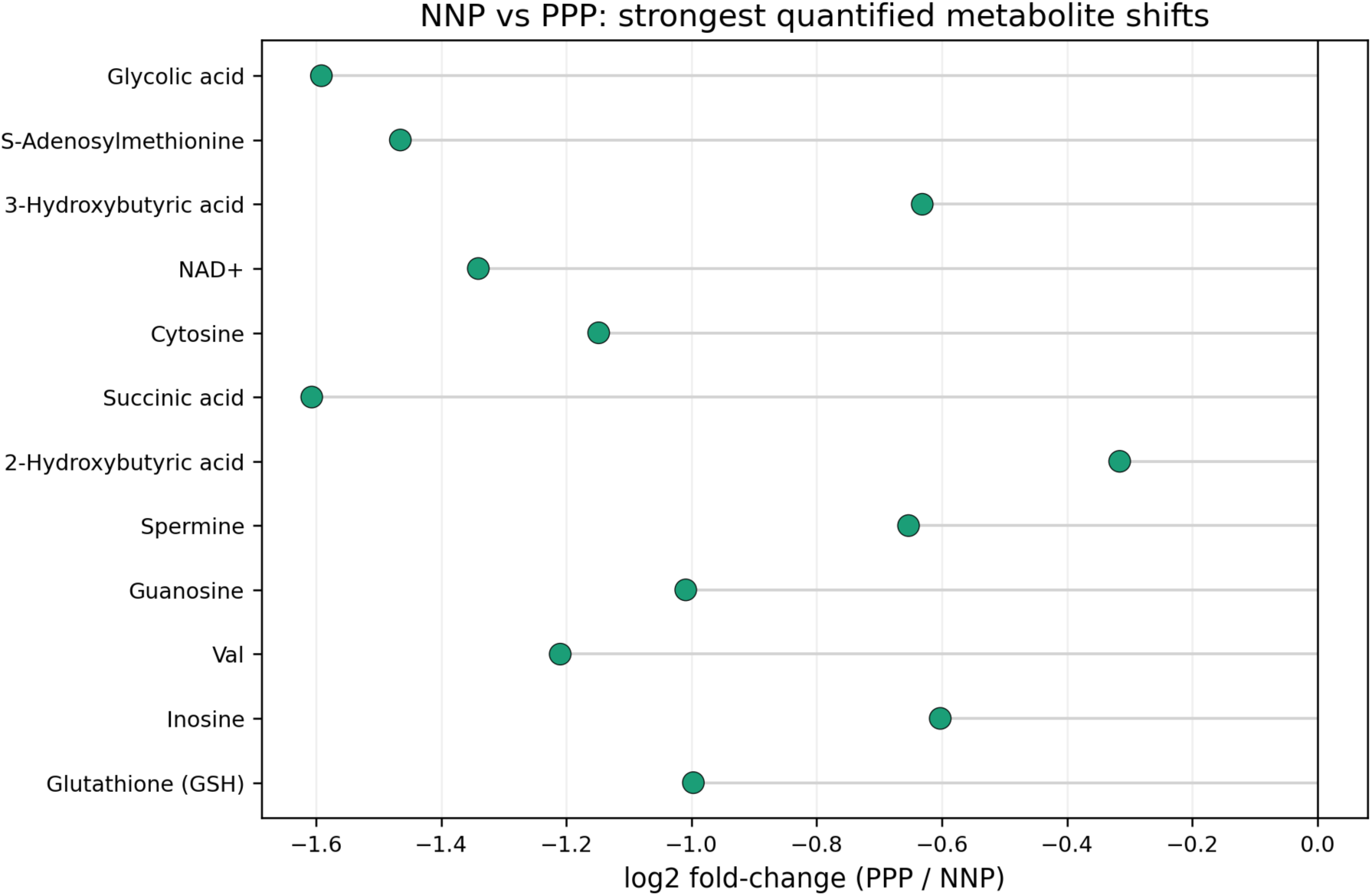
Quantitative metabolites most strongly shifted between NNP and PPP.

**Fig. 9.**
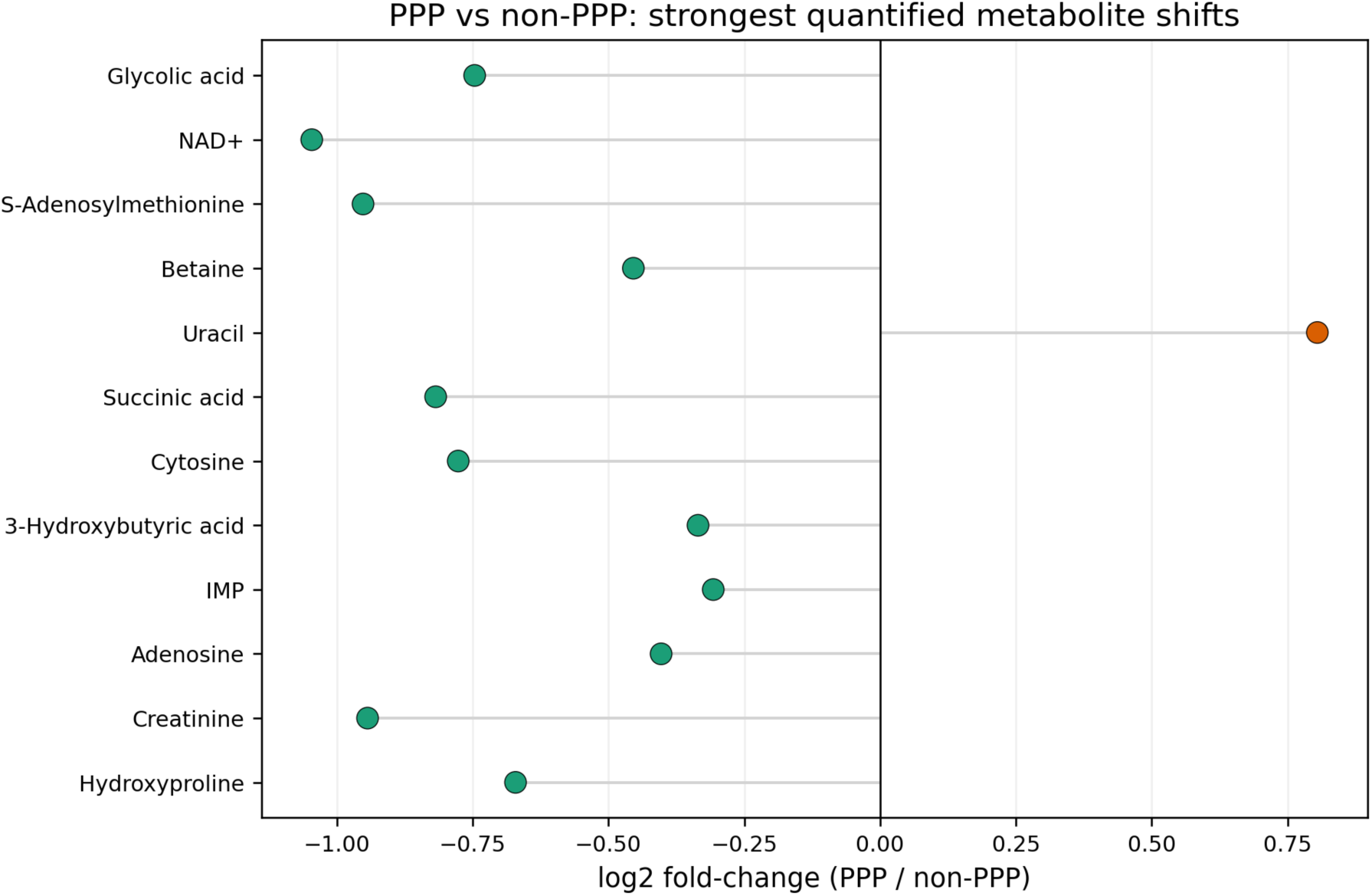
Quantitative metabolites contributing to PPP versus non-PPP discrimination.

### Metabolite ratios expose latent transition-state information

Ratio mining improved discrimination in several biologically relevant contrasts (Figure 10). For PPP versus non-PPP, uracil/S-adenosylmethionine and uracil/creatinine each achieved ROC AUC values of 0.775, followed closely by uracil/glycolic acid (AUC 0.764). These ratios suggest that persistent positivity is better represented by the balance between pyrimidine-related signal and methyl-donor or housekeeping metabolites than by any one feature alone. For NNP versus PPP, the glycolic acid/glutamine ratio reached an AUC of 0.872 (Figure 11). For NNP versus NPP, glucose 1-phosphate/isovaleric acid-valeric acid produced an AUC of 0.897, indicating that metabolic ratios can sharpen the separation between different HPV transition paths even when global class structure is weak (Figure 12).

**Fig. 10.**
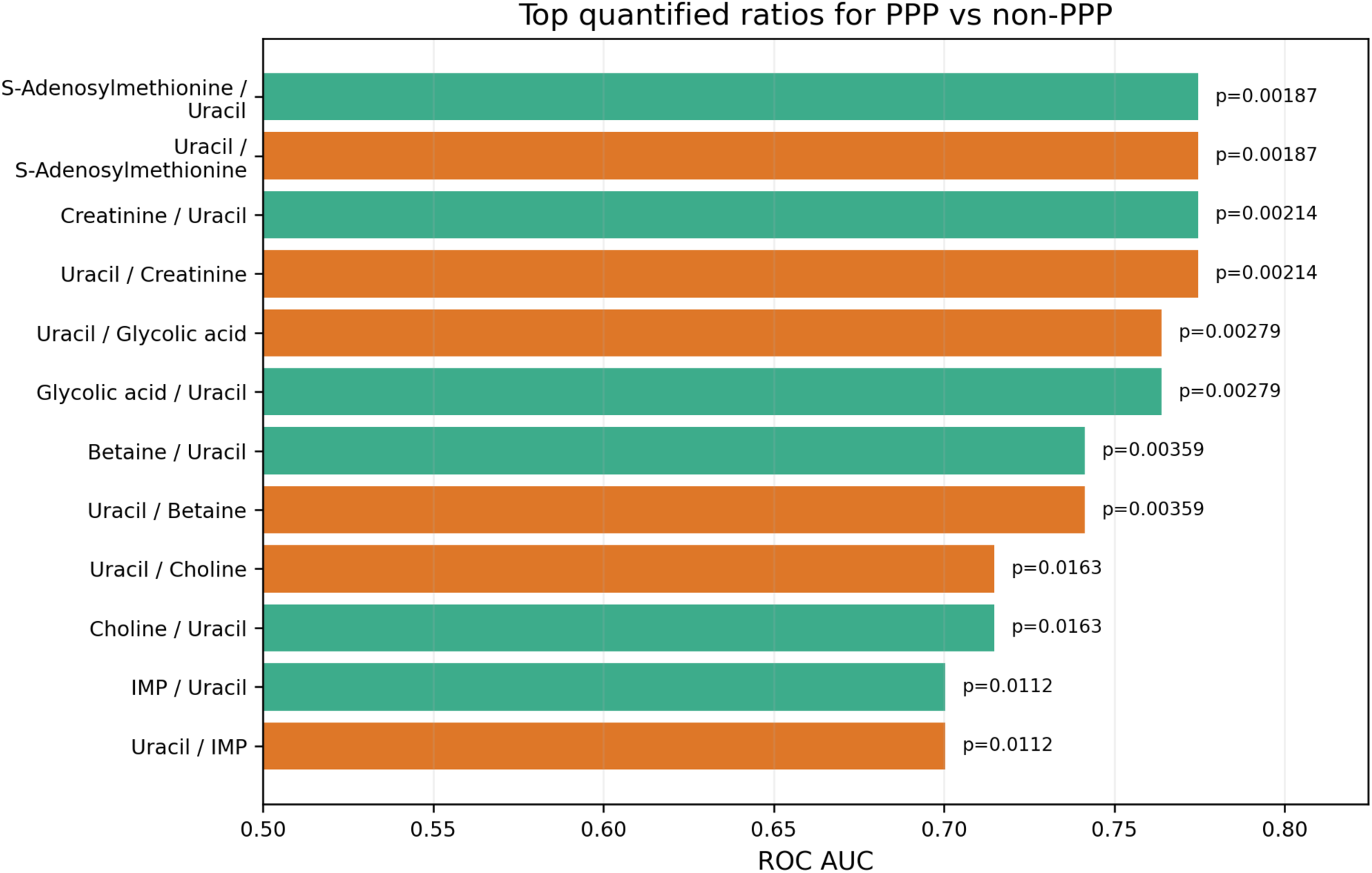
Top metabolite ratios for PPP versus non-PPP.

**Fig. 11.**
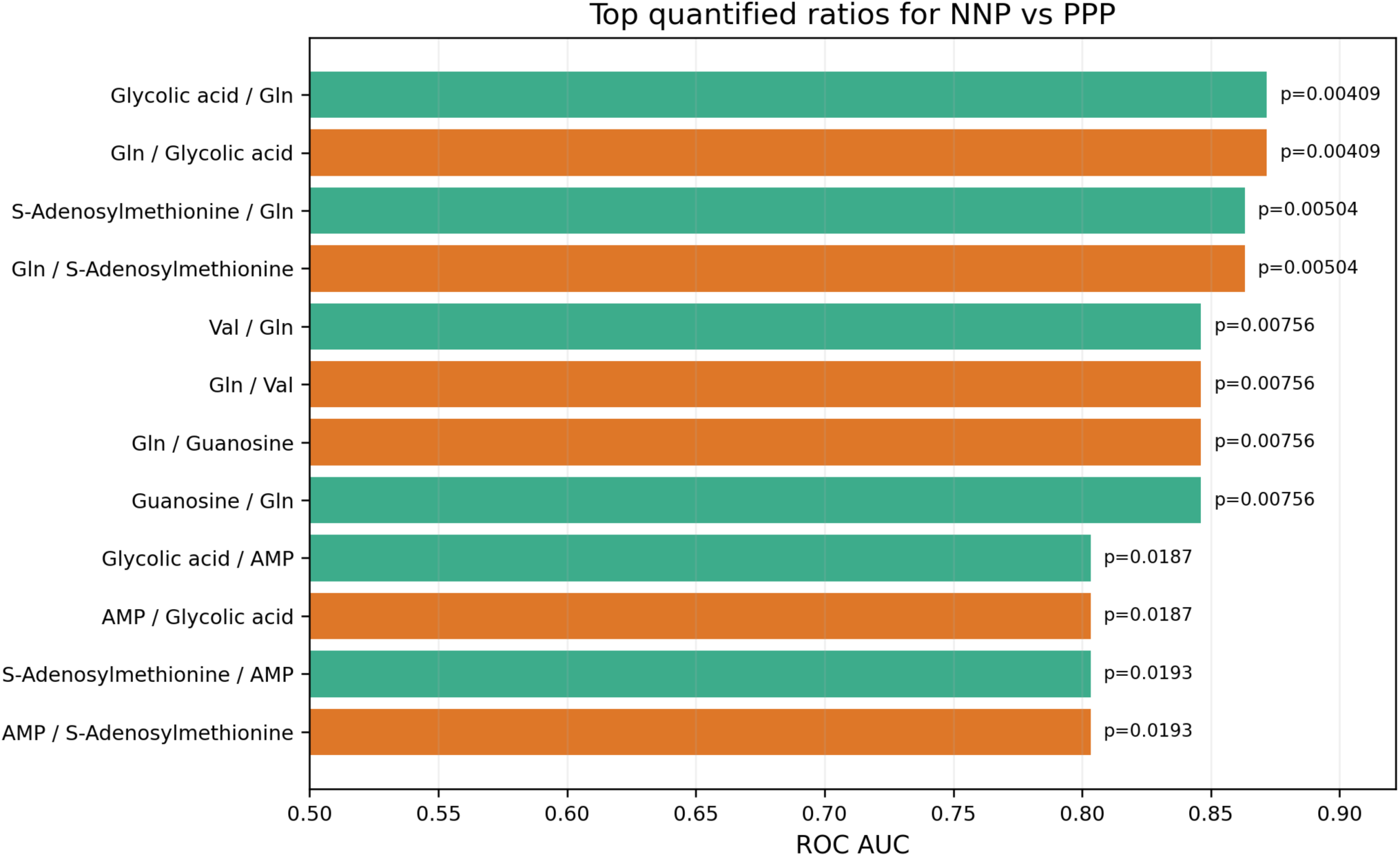
Top metabolite ratios for NNP versus NPP.

**Fig. 12.**
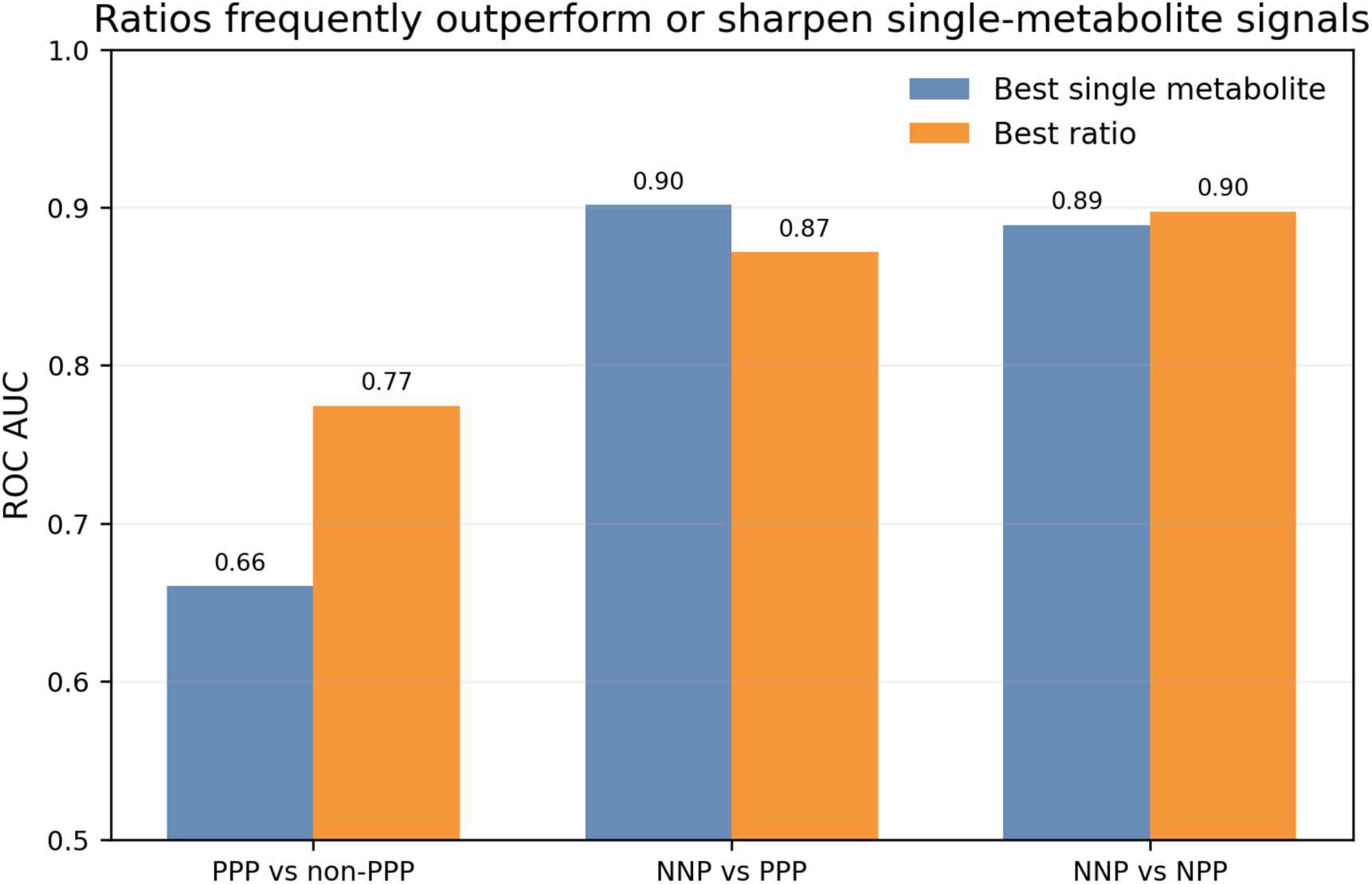
Ratio features often sharpen contrast performance relative to the best single metabolite.

### Module, pathway, and network analyses suggest focused rather than diffuse remodeling

Module-level analyses provided only modest additional evidence. Modules M1 and M2 were nominally different across classes, but no module survived global false-discovery correction, reinforcing the conclusion that large coordinated network shifts are limited Figure 13). Pathway analyses were more informative when restricted to the PPP-focused comparisons. In PPP versus non-PPP enrichment, carbon fixation pathways in prokaryotes, glyoxylate and dicarboxylate metabolism, and the tricarboxylic acid cycle were enriched, driven principally by succinic acid, citric acid, malic acid, and glycolic acid (Figure 14). These results place glycolic acid in a broader dicarboxylate- and central-carbon-linked context.

**Figure 13:**
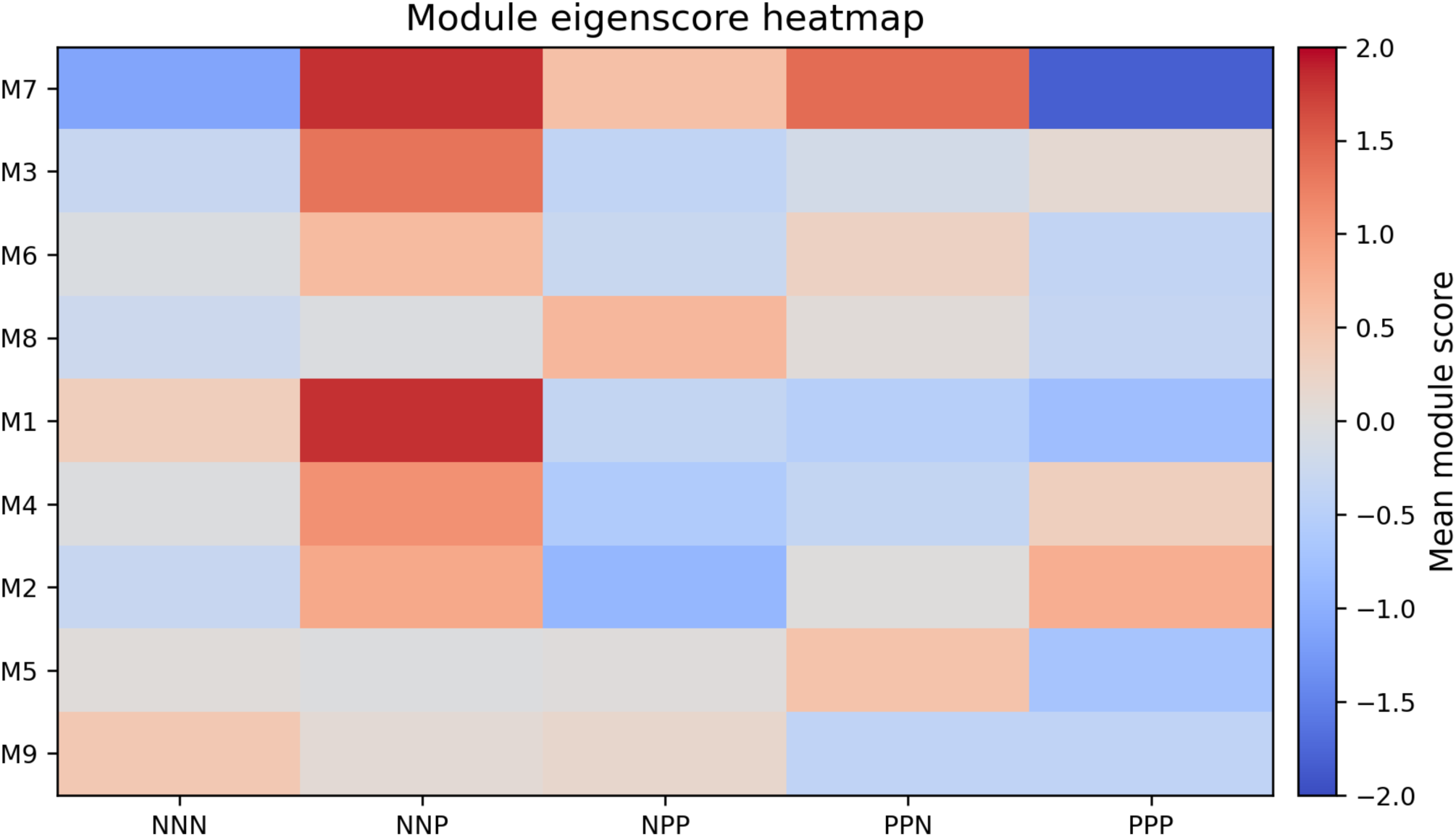
Heatmap of means of module groups by class trajectory.

**Fig. 14.**
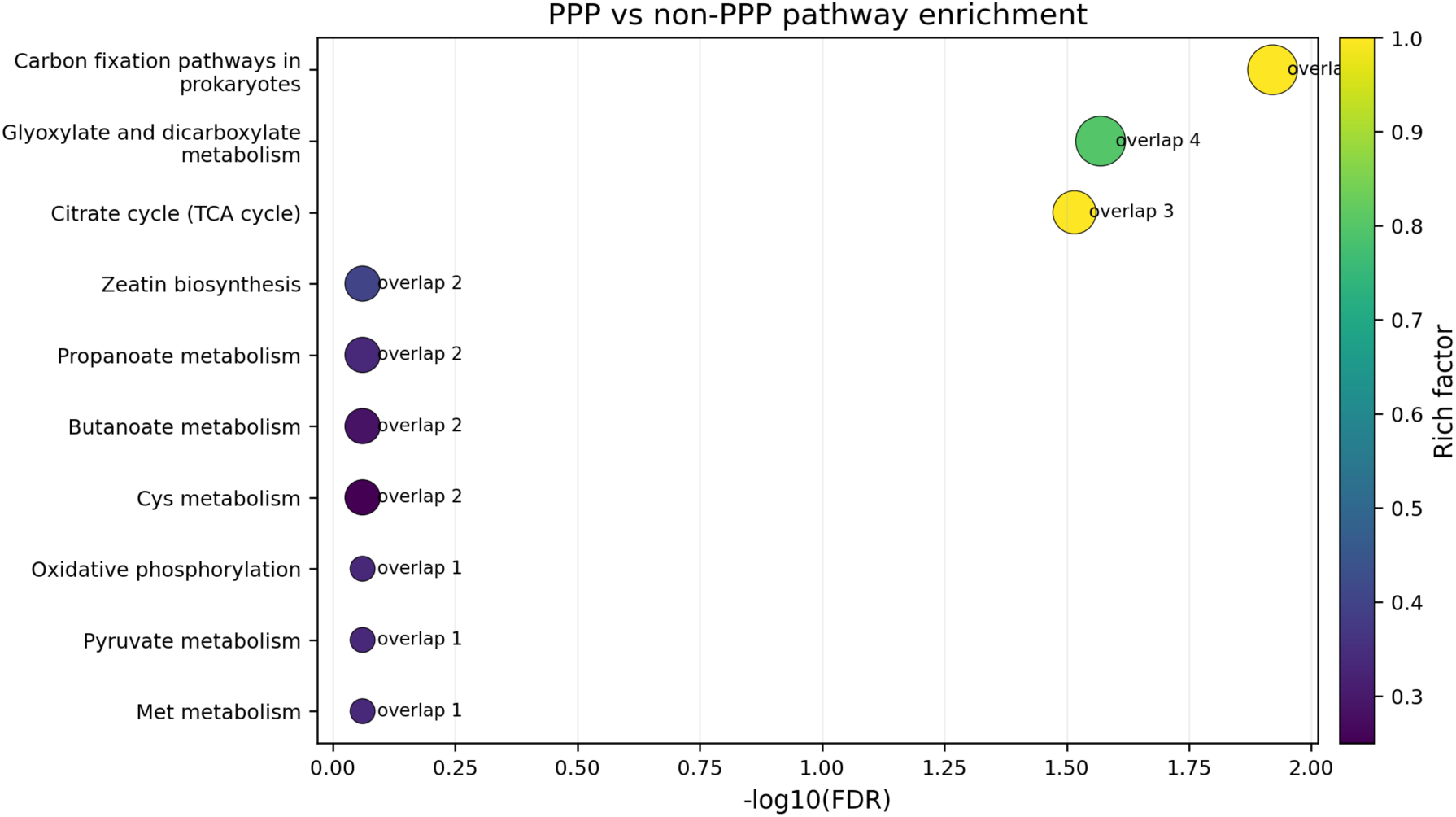
Pathway enrichment for PPP versus non-PPP indicates a central-carbon/dicarboxylate theme.

A complementary network view showed that the strongest quantified NNP-versus-PPP metabolites also underwent marked correlation rewiring. Together with SHAP and PLS-VIP analyses, these results converged on glycolic acid, S-adenosylmethionine, NAD+, betaine, uracil, and related metabolites as the most recurrent quantitative markers (Figure 15, Supplementary Figure 7). By contrast, trajectory encoding using burden or slope produced only weak associations after correction, consistent with class-specific metabolic states dominating over simple monotonic “more HPV burden equals more metabolic disruption” models (Figure 16). Figure 17 shows a working model of the HPV trajectory encoded by targeted metabolic states and the four interconnected pathway clusters.

**Fig. 15.**
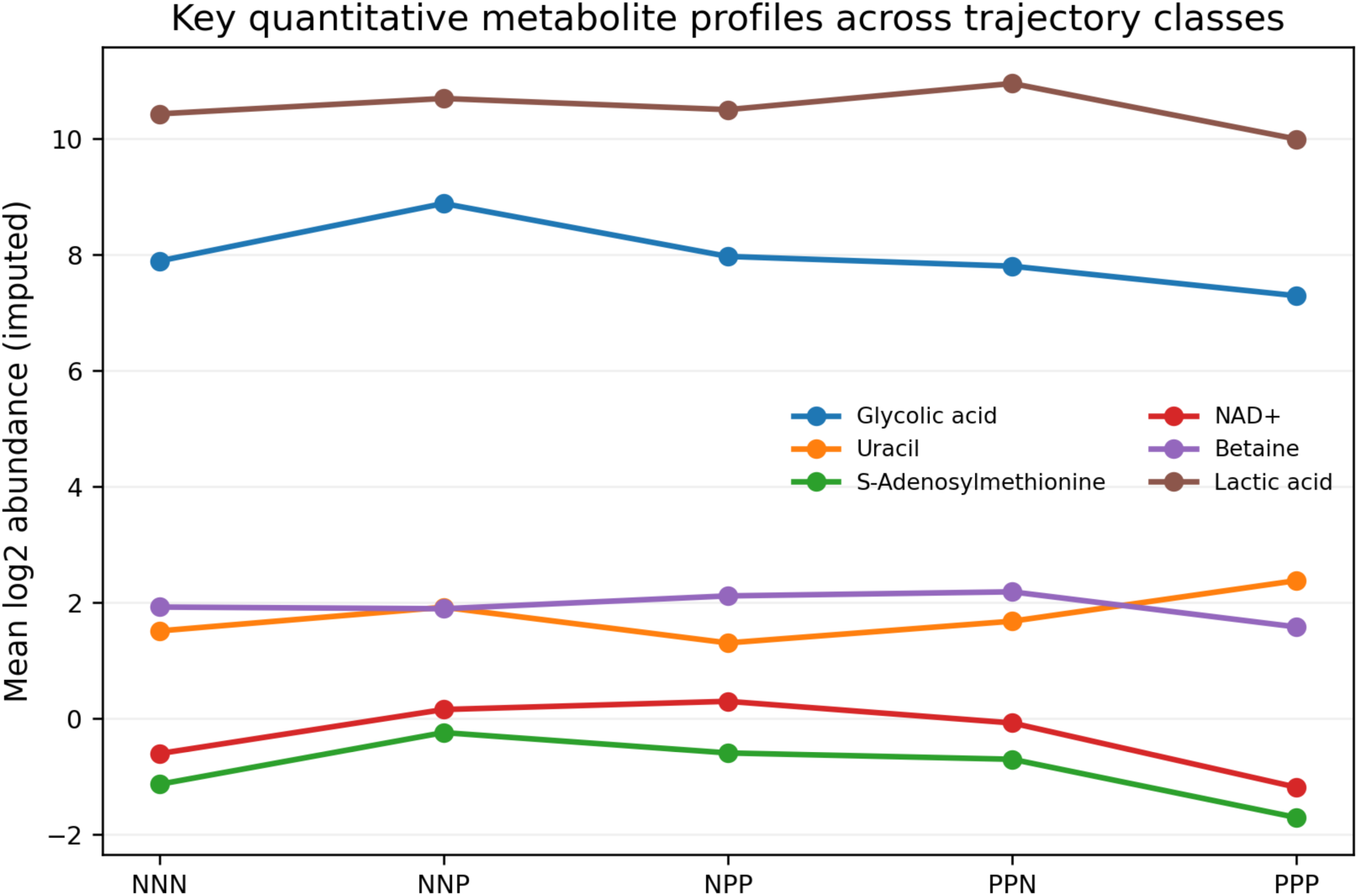
Profiles of representative quantified metabolites across HPV trajectory classes.

**Fig. 16:**
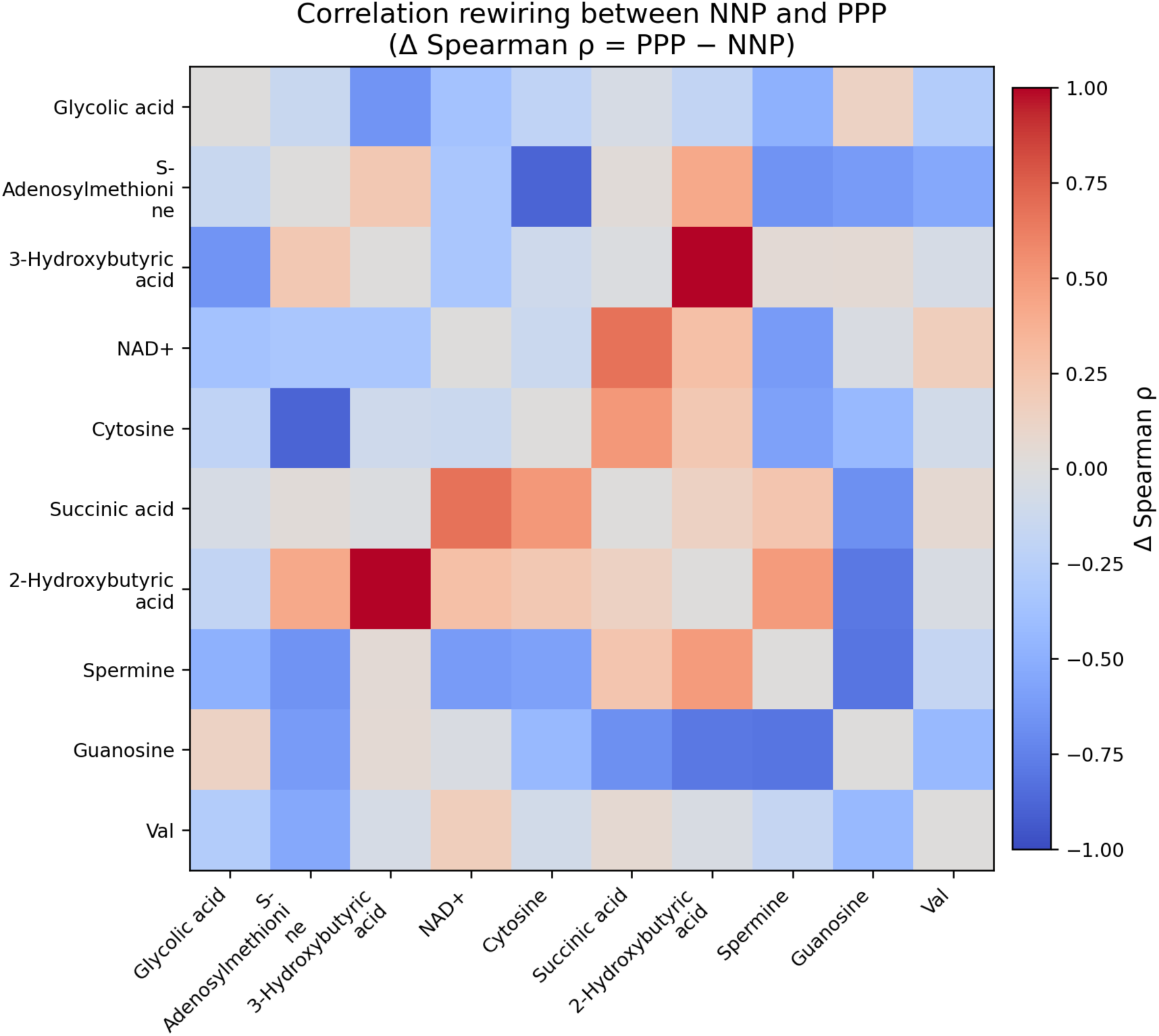
Correlation rewiring among leading quantitative markers between NNP and PPP.

**Fig. 17.**
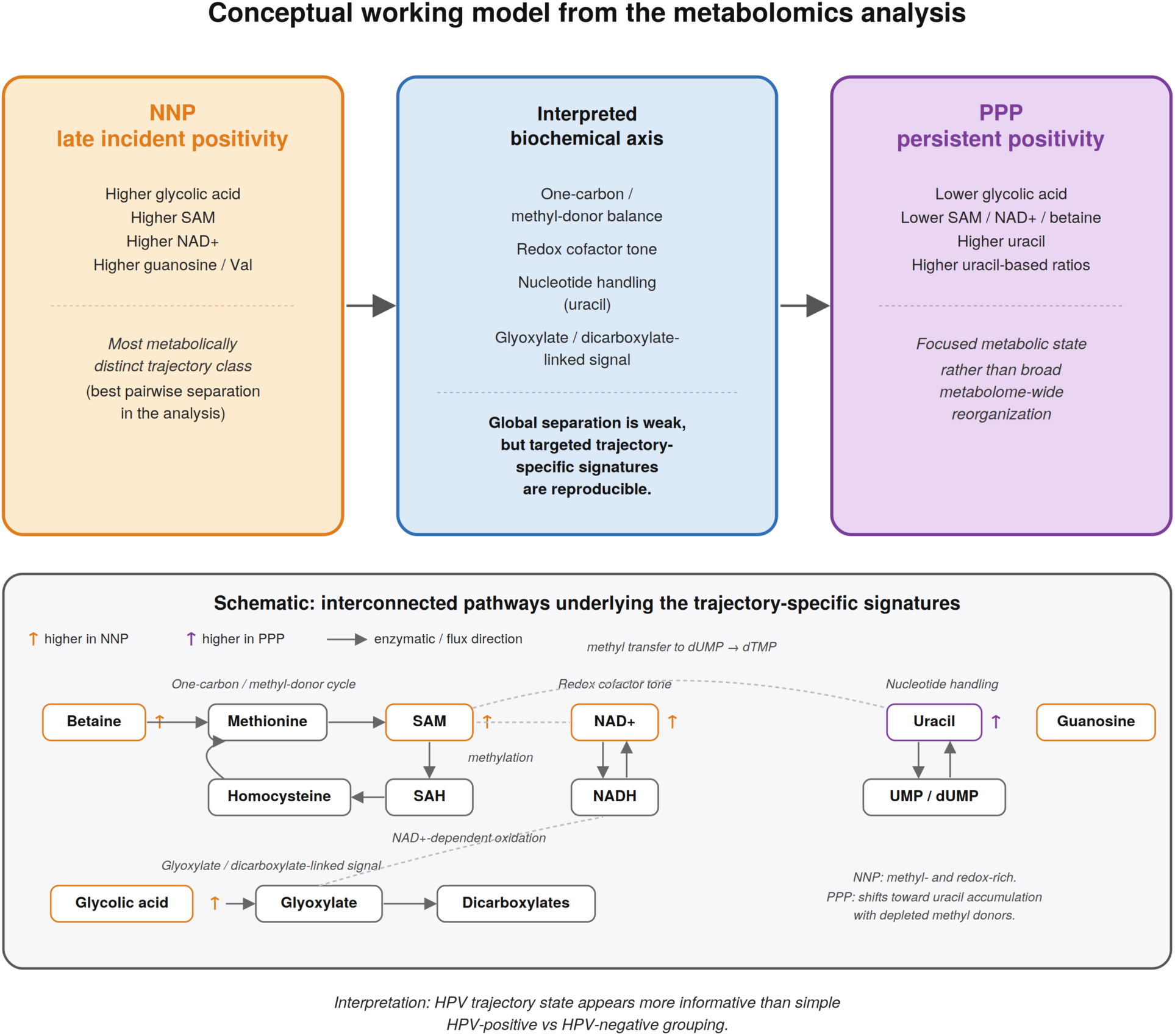
Working model: HPV trajectory is encoded by targeted metabolic states rather than a broad positive-versus-negative shift.

## Discussion

The principal conclusion of this study is that the vaginal metabolome in this tightly phenotyped HIV-negative cohort does not strongly separate all HPV trajectory classes at a global level, however, when HPV is modeled as a time-resolved trajectory rather than a binary exposure, reproducible and biologically interpretable signatures emerge - most clearly for late incident positivity (NNP) and persistent positivity (PPP). This distinction is important because a conventional HPV-positive versus HPV-negative framing would make the dataset appear largely “negative,” whereas a trajectory-aware approach reveals focused metabolic states that plausibly reflect transition and persistence biology. These patterns are consistent with prior cervicovaginal metabolic profiling work in which HPV-related functional signals are embedded within broader host–microbe and inflammatory variation and become clearer when modeled with disease/host context rather than as a single global axis.^21,22^

The most striking result was the prominence of glycolic acid. Glycolic acid emerged as the dominant single metabolite for NNP versus PPP, showed convergent evidence from both abundance and detection, and also recurred across multiple analytic layers, whoever its mechanistic source cannot be assigned with confidence because we did not include microbiome, pH, or epithelial transcriptomic data in this analysis. Nevertheless, its repeated co-appearance with dicarboxylate- and central-carbon-related pathway signals suggests that it indexes a broader biochemical context involving glyoxylate/dicarboxylate handling or linked host-microbial carbon processing rather than a measurement artifact.

The PPP phenotype was notable not for scale but for coherence. Relative to NNP and non-PPP states, persistent positivity was characterized by lower S-adenosylmethionine, lower NAD+, lower betaine, and higher uracil. This combination suggests a state in which methyl-donor availability, redox tone, and nucleotide handling are altered in concert. Such a pattern is plausible in the setting of persistent viral presence, where epithelial turnover, nucleotide demand, and host response may remain chronically engaged even in the absence of a large global metabolomic displacement. Notably, integrated microbiota–metabolome analyses of persistent HPV infection accompanied by high-grade CIN have reported extensive metabolite shifts and have explicitly proposed bi-directional VM–HPV modulation that may be partially mediated by vaginal metabolites, supporting the general plausibility of a persistence-associated “functional” state even when compositional shifts are modest^23^.

The uracil-centered ratios are particularly informative. Uracil/S-adenosylmethionine, uracil/creatinine, and uracil/glycolic acid all performed better than most single metabolites for PPP classification. Ratios often capture pathway imbalance more effectively than absolute levels, particularly in small discovery datasets where dilution effects, intermittent non-detection, and broad between-subject variation can obscure biologically meaningful contrasts. In the present study, ratio mining exposed latent structure that standard global analyses largely missed.

An additional insight is that NNP was more distinctive than PPP in many analyses. This late incident positivity state—negative, negative, then positive—may represent a sharper transition than chronic positivity itself. One interpretation is that incident positivity is accompanied by a temporally narrow biochemical window marked by higher glycolic acid and related metabolites, whereas persistent positivity settles into a more stable but lower-amplitude metabolic state. This interpretation is consistent with the class geometry: NNP is the most distant centroid, whereas NNN and PPP are unexpectedly close. This transition sensitivity also aligns with clinical-transition datasets in which metabolomic profiles have been reported to respond strongly to changed HPV infection state and, in some contexts, to outperform microbiota composition for describing vaginal microenvironment response^24^.

This subtlety helps reconcile our findings with the broader cervicovaginal literature. Vaginal microbial community state strongly influences metabolites linked to lactic acid biology, carbon substrate availability, and inflammation^4-10,14,19,20,25^. In more heterogeneous cohorts, HPV detection and cervical disease are often accompanied by broad ecological differences, which can dominate metabolomic structure and make HPV-associated signals appear either dramatic (when dysbiosis co-varies with HPV) or inconsistent (when background heterogeneity is high).^6,7,15,17,18,26^ In contrast, the current cohort was deliberately constrained: HIV negative, no other STI, premenopausal, non-smoking, and batch-matched. Within this cleaner background, the metabolomic footprint of HPV appears subtler, more state-specific, and not reducible to a classic bacterial vaginosis-like signature. Cross-sectional vaginal metabolomics work has nevertheless demonstrated HPV-associated differences that persist after accounting for bacterial community state type, underscoring that HPV status can be associated with functional shifts even when dominance patterns are similar^27^. Mechanistically, HPV may also actively reshape the cervicovaginal microenvironment by altering host mucosal innate peptide expression in ways that affect Lactobacillus nutrient access, providing a plausible chain from HPV infection to host peptide changes to microbiota shifts unto metabolite remodeling, which is compatible with a focused metabolic imprint and without requiring a wholesale BV-like transition^28^.

A useful way to reconcile our findings with an alternative PCA-based analyses is to distinguish metabolites that dominate unsupervised variance from those that reproducibly discriminate HPV trajectory. In the PCA, lactic acid loads strongly on PC1, and PC2 is influenced by SCFA- and keto-acid-related features such as γ-butyrobetaine, propionic acid, pyruvic acid, and the isobutyric acid/butyric acid signal; however, PC1 and PC2 themselves were not associated with trajectory class, lactic acid was detected in all samples and showed no global class difference, and the low-abundance SCFA/keto-acid features were either weakly class-associated or too sparse for stable inference after filtering and multiple-testing correction. The same principle applied to several pathway-linked metabolites highlighted in the enrichment analyses. Citric acid, malic acid, succinic acid, and 3-hydroxypropionic acid helped define the PPP-associated carbon fixation, glyoxylate/dicarboxylate, and TCA themes, yet none emerged as a robust standalone HPV marker after correction, consistent with coordinated low-amplitude pathway shifts rather than a single dominant discriminator. Likewise, γ-butyrobetaine and the isovaleric acid/valeric acid feature captured biologically plausible background or contrast-specific structure, but their effects were restricted to particular pairwise comparisons and remained sensitive to sparsity, detection heterogeneity, or feature ambiguity. By contrast, S-adenosylmethionine, NAD+, betaine, and uracil illustrate the reverse situation: they were not dominant drivers of global PCA variance yet became informative once the analysis was aligned to persistent-positivity contrasts and ratiometric imbalance. Taken together, these data indicate that the largest latent axes in our study are not synonymous with HPV-associated variation; the most reproducible HPV signal in this cohort resides in focused trajectory-specific contrasts and metabolite ratios rather than in the metabolites with the highest unsupervised loadings.

Importantly, targeted interrogation of keto acids, methylated nucleosides, and pathway-level signatures did not support a narrative of broad metabolite accumulation across all HPV-positive women. Neither 2-hydroxybutyric acid nor 3-hydroxybutyric acid showed global enrichment by trajectory class, and both were too sparse or too contrast-specific to justify presentation as major markers of the HPV-positive microenvironment. The same caution applied to RNA-modification- and methylation-related features such as 1-methyladenosine, 1-methyl-4-imidazoleacetic acid, and 1-methyl-2-pyrrolidone: these metabolites were detected in a minority of samples and did not yield stable, corrected associations, indicating that they do not provide a robust secondary predictive layer for progression from incident to persistent infection in this dataset. More informative than these sparse methylated analytes were the coordinated changes observed in uracil, S-adenosylmethionine, NAD+, and betaine, particularly in persistent positivity, which point to a focused disturbance in pyrimidine balance, methyl-donor availability, and redox metabolism rather than to generalized accumulation of modified purines or imidazoles. This “focused remodeling” framing is compatible with adjacent cervicovaginal metabolomics work in high-risk HPV infection stratified by lesion severity, where discriminative metabolite panels can differ substantially by sampling matrix and disease spectrum (e.g., NILM vs LSIL vs HSIL), reinforcing that HPV-associated metabolic signals are often state-dependent rather than universal^29^.

The study has several strengths. The cohort is unusually homogeneous with respect to major biological and behavioral confounders. All samples were collected in the same broad time frame and processed together, limiting batch variation. The analysis also moved beyond simple differential abundance testing by integrating detection prevalence, machine learning, permutation testing, ratio mining, module/pathway scoring, and feature recurrence. The resulting conclusions are therefore grounded in convergence across multiple analytic layers rather than any single *p* value.

The study also has several limitations. First, the sample size is modest, especially for NNP, so all machine-learning results should be interpreted as discovery-stage rather than clinical-grade classifiers. Second, we did not adjust for participant-level hormonal-cycle data, contraceptive exposure, vaginal pH, sexual behavior variables, type-specific HPV genotype information, or HPV viral-load measures. Recent synthesis has emphasized that cervicovaginal microbiome–HPV studies must “catch up with biology” by incorporating richer HPV metrics (e.g., genotype-resolved persistence biology and mechanistic endpoints) to avoid overinterpreting community-level associations^30^. Third, the study is trajectory-defined but not truly serial at the metabolite level: each woman contributes one metabolomic snapshot that is interpreted against longitudinal HPV status rather than repeated within-person metabolomics over time. Fourth, pathway annotations are partly database-derived and should be treated as directional rather than definitive mechanistic proof. Fifth, missing values were interpreted as non-detection and imputed for some analyses. While this is pragmatic, left-censoring and batch/signal-drift issues in untargeted MS can materially affect low-abundance features, underscoring the importance of explicitly reporting system suitability, pooled QC strategies, and signal stability and aligning QA/QC reporting with current consortium guidance for untargeted metabolic phenotyping^31,32^. Because metabolomic profiling was performed at a single sampling time and no parallel microbiome, vaginal pH, or Nugent-score data were available, our analyses cannot establish true temporal metabolite dynamics, direct vaginal dysbiosis, or causal SCFA-mediated toxicity.

Taken together, the findings support a restrained but biologically interesting model. High-risk HPV does not impose a strong universal vaginal metabolome signature in this healthy HIV-negative population. Instead, specific transition states—especially late incident positivity and persistent positivity—leave a focused metabolic imprint centered on glycolic acid and on a PPP-associated uracil-high, methyl-donor/redox-low state. This trajectory-centric interpretation is also consistent with clinical-transition studies in which cervicovaginal microbiome and metabolome profiles shift alongside HPV clearance after excisional treatment and then continue to evolve over 6–12 months as the vaginal microenvironment restores^33,34^. Future work should test whether this metabolite constellation replicates in larger cohorts, maps onto vaginal microbiome state, HPV genotype and activity measures, and local immune mediators, and predicts persistence versus clearance prospectively.

## Methods

### Cohort definition

Participants were women enrolled in the ACCME Study who volunteered for more intensive vaginal sampling every 6 months at baseline, 6 months, and one-year in 2019 and 2021^35^. A total of 100 participants in the following five categories based on their HPV test results were invited to participate. The five categories are:

1. 20 women who are persistently negative for high-risk HPV infection (hrHPV---) = NNN
2. 20 women who may be newly infected after two consecutive negative tests (hrHPV--+) = NNP
3. 20 women who may be newly infected after one negative test for high-risk HPV (hrHPV-++) = NPP
4. 20 women whose status changed after 2 positive tests (hrHPV++-) = PPN
5. 20 women who are persistently positive for high-risk HPV infection (hrHPV+++) = PPP

These women were followed up for 12 weeks and two mid-vaginal samples collected twice a week from them, prospectively. Only 71 participants who provided analyzable samples were enrolled in the study. All participants were self-reported HIV negative, free of other sexually transmitted infections after pre-enrollment testing (high vaginal swab and urine microscopy, culture, and sensitivity, and serum samples for *C. trachomatis* and *T. pallidum*), premenopausal, non-smoking, and sampled within a common laboratory workflow. The mean age of the cohort was 39 years, and all participants had history of sexual activity.

### Metabolic analysis

All samples were shipped to Human Metabolomics Technologies (HMT) from Nigeria in -80°C shippers and temperature was maintained throughout the shipment verified by shipment logs. At HMT, 20 μL of samples were mixed with 20 μL of Milli-Q water containing internal standards (1,000 μM) and 60 μL of Milli-Q water. The compounds were measured in the Cation and Anion modes of CE-TOFMS based metabolome analysis in the standard conditions^36-38^. Peaks detected in CE-TOFMS analysis were extracted using automatic integration software (MasterHands ver. 2.17.1.11 developed at Keio University) in order to obtain peak information including *m/z*, migration time (MT), and peak area. The peak area was then converted to relative peak area by the following equation.

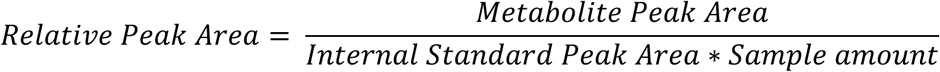

The peak detection limit was determined based on signal-noise ratio; S/N = 3.

Putative metabolites were then assigned from HMT’s standard library and Known-Unknown peak library on the basis of *m/z* and MT. The tolerance was ±0.5 min in MT and ±10 ppm in m/z.

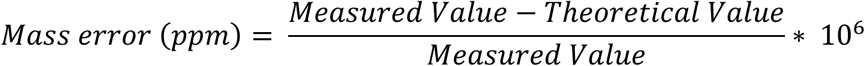

If several peaks were assigned the same candidate, the candidate was given the branch number.

Absolute quantification was performed in target metabolites. All the metabolite concentrations were calculated by normalizing the peak area of each metabolite with respect to the area of the internal standard and by using standard curves, which were obtained by single-point (100 *μ*M or 50 *μ*M) calibrations

#### Metabolite matrices

The data consist of two principal analytical layers: putative metabolites measured as relative area and a targeted quantitative layer measured as estimated concentrations. Non-detected values were treated as missing. Downstream analyses were performed on the validated filtered feature sets contained in the companion analysis workbook, comprising 186 putative and 64 quantified metabolites. These filtered sets were extracted from the raw spreadsheet to regenerate visualizations and manuscript figures.

#### Preprocessing

Missing values were interpreted as non-detection events. For analyses requiring continuous matrices, missing values were imputed feature-wise using one-half of the minimum positive observed value for that feature, followed by log2 transformation. For multivariate visualization, machine learning, network analysis, and pathway/module scoring, features were standardized to z scores across samples. Detection prevalence was retained separately to permit two-part analyses combining abundance and detection evidence.

#### Univariate statistical testing

Global multi-class testing used Kruskal–Wallis and analysis-of-variance summaries as captured in the validated workbook. Pairwise and grouped binary contrasts were assessed with Mann–Whitney tests together with effect size estimates, fold-change summaries, and receiver operating characteristic area under the curve (ROC AUC). *p-*values were adjusted by false discovery rate where appropriate. Detection prevalence contrasts were evaluated with contingency-based tests. For selected key comparisons, abundance and detection *p* values were combined to prioritize metabolites supported by both dimensions.

#### Multivariate and machine-learning analysis

Principal component analysis was used for unsupervised visualization. Supervised discrimination was evaluated with elastic-net logistic regression, linear support vector machines, and random forests. Performance was summarized using balanced accuracy and ROC AUC under repeated stratified cross-validation in the validated analysis workbook. One-versus-rest and grouped contrasts were analyzed in addition to exhaustive pairwise comparisons. Permutation testing was used to assess whether the strongest observed pairwise signals exceeded chance expectation.

#### Stability selection, SHAP, and PLS-VIP

Feature recurrence across resampled penalized models was summarized as stability selection frequency. For interpretability of the strongest quantitative pairwise contrast (NNP versus PPP), a random forest model was fitted and TreeSHAP mean absolute importance values were computed. Partial least-squares discriminant analysis was used in an exploratory manner to generate variable importance in projection (VIP) rankings for NNP versus PPP and PPP versus non-PPP.

#### Ratio mining

Pairwise ratios were screened among selected quantified metabolites to identify compound balances that improved discrimination beyond individual markers. Ratios were ranked by ROC AUC and univariate significance. This step was motivated by the observation that trajectory-associated metabolite imbalances may be more stable than absolute abundance differences in small cohorts.

#### Module, pathway, and network analyses

Putative metabolites were grouped into correlation-derived modules in the validated workbook, and module scores were summarized across classes. Pathway enrichment was performed on prioritized metabolite sets, and representative pathway scores were visualized as the mean standardized abundance of pathway-member metabolites. Correlation rewiring was examined by comparing within-class Spearman correlation structures, particularly between NNP and PPP for leading quantitative metabolites. Recurrent markers were defined as metabolites that reappeared across multiple analytic layers, including pairwise testing, two-part evidence, stability selection, and curated selected-feature sets.

#### Trajectory encoding

In addition to categorical class analysis, trajectory burden and slope encodings were explored to ask whether increasing HPV exposure or temporal direction generated monotonic metabolite trends. These regression-based analyses were weak after multiple-testing correction, supporting the interpretation that class-specific states dominated over simple scalar encodings of HPV burden.

#### Software and outputs

Final visualizations, the manuscript document, and the downloadable figure archive were generated using Python. The companion Excel workbook contains the full tabulated results used to construct the present manuscript and figure set.

## Data availability

Source data for this analysis are available on the ACCME Project websites https://h3accme.org/

## Acknowledgments

The project described was supported by the African Collaborative Center for Microbiome and Genomics Research grants (NHGRI/NIH U54HG006947) from the NIH Office of the Director/NHGRI, Maryland Department of Health Cigarette Restitution Fund Program (grant number CH-649-CRF), and the University of Maryland Greenebaum Comprehensive Cancer Center Support Grant (National Cancer Institute Award Number: P30CA134274). The funding agencies did not play any role in data collection, analysis, or publication.

We sincerely thank Dr. Alex Buko of Human Metabolome Technologies for analysis of samples and data. We thank all participants and researchers in the ACCME Research Project.

## Author information

CAA developed, designed it, obtained funding, supervised implementation, data management and analysis of the study, and wrote the first draft. SNA contributed to the study design, data management and analysis. GT, OI, YO, and AF enrolled study participants, collected and managed local storage of samples, and conducted screening tests. All authors contributed to drafting the manuscript, provided critical revisions and approved the final draft.

## Ethics declaration

Ethical approval was obtained from the National Health Research Ethics Committee of Nigeria (NHREC/01/01/2007-26/10/2025I) and the institutional ethics committees at the University of Maryland School of Medicine, Baltimore (US) (IRB Number HP-00058830). All participants gave written informed consent in accordance with the Declaration of Helsinki and the Nigerian National Code for Health Research Ethics.

## Supplementary images

**Supplementary Figure 1:**
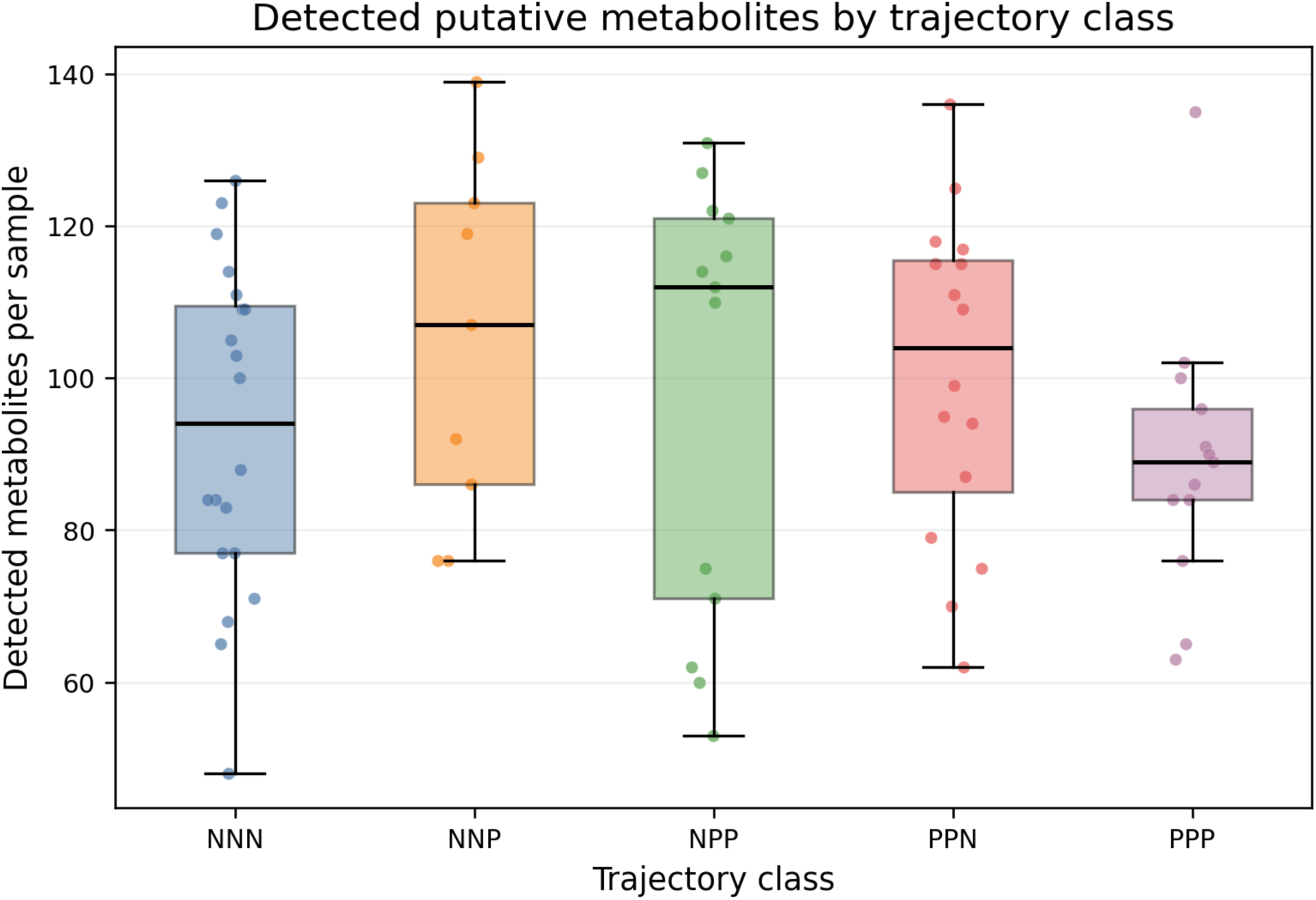
Detection burden of putative metabolites’ trajectory across classes.

**Supplementary Figure 2:**
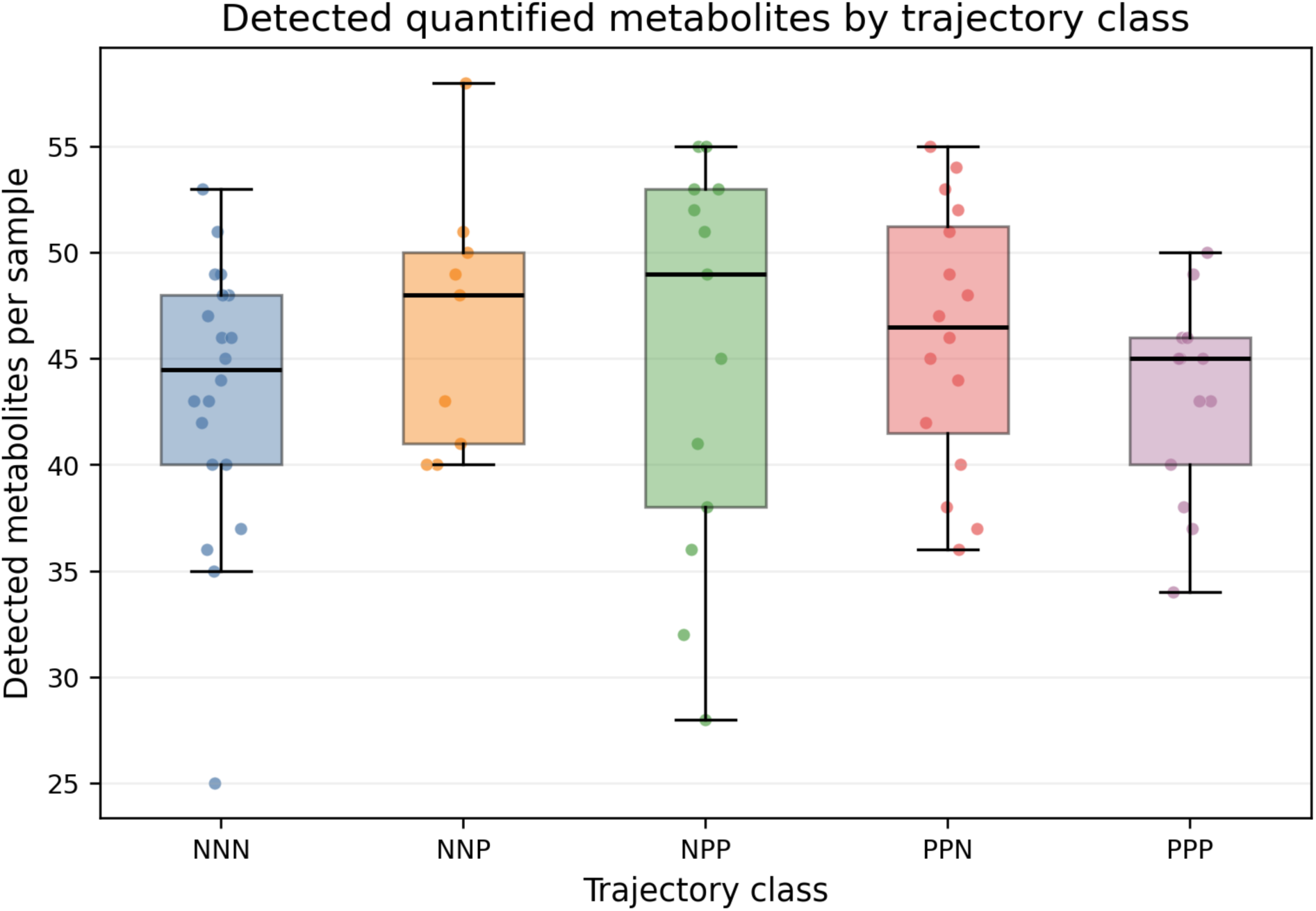
Detection burden of quantified metabolites’ trajectory across classes.

**Supplementary Figure 3:**
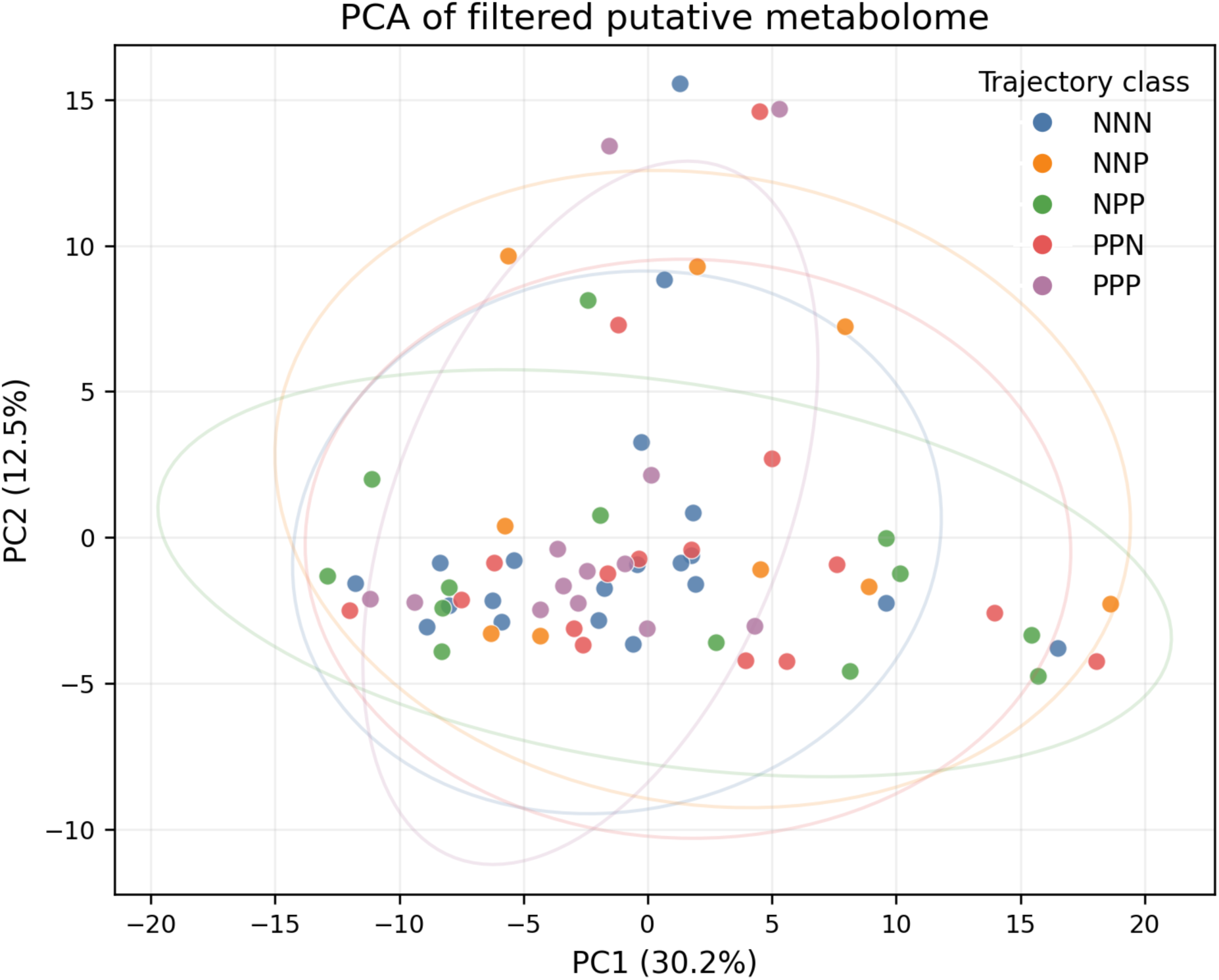
Principal component analysis of both the putative metabolome matrices.

**Supplementary Figure 4:**
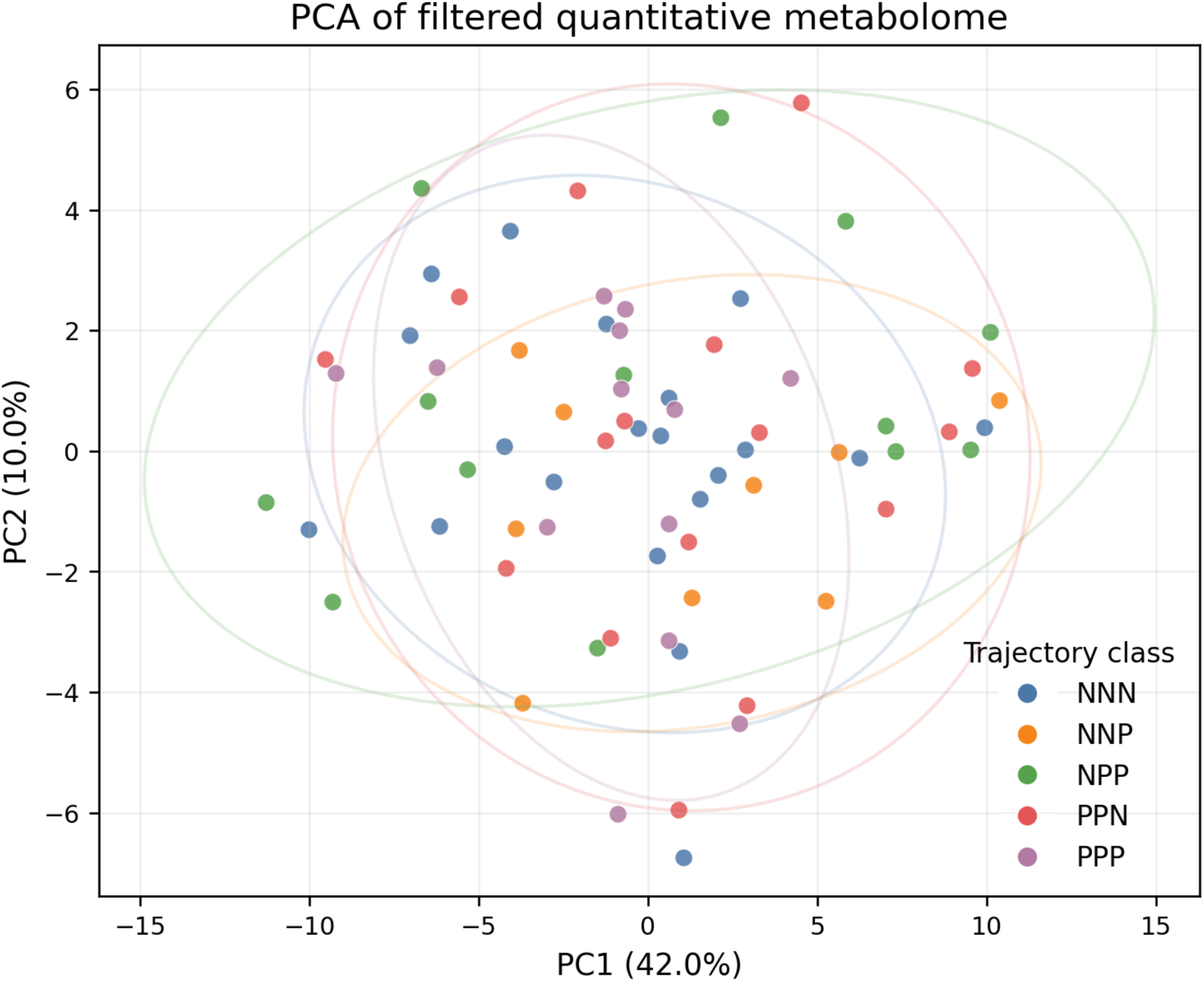
Principal component analysis of quantitative metabolome matrices.

**Supplementary Figure 5:**
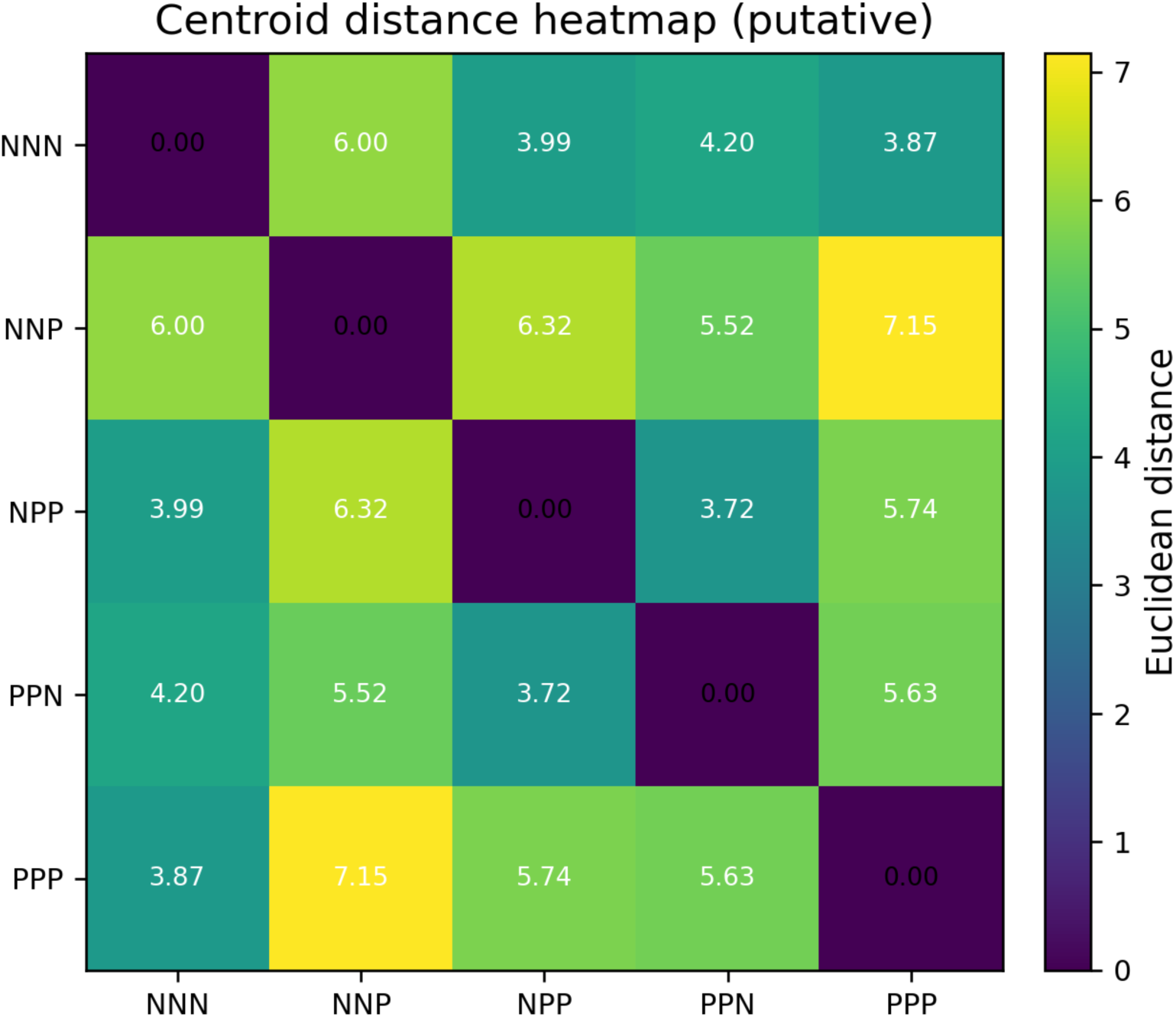
Quantitative centroid distances of putative metabolites across classes.

**Supplementary Figure 6:**
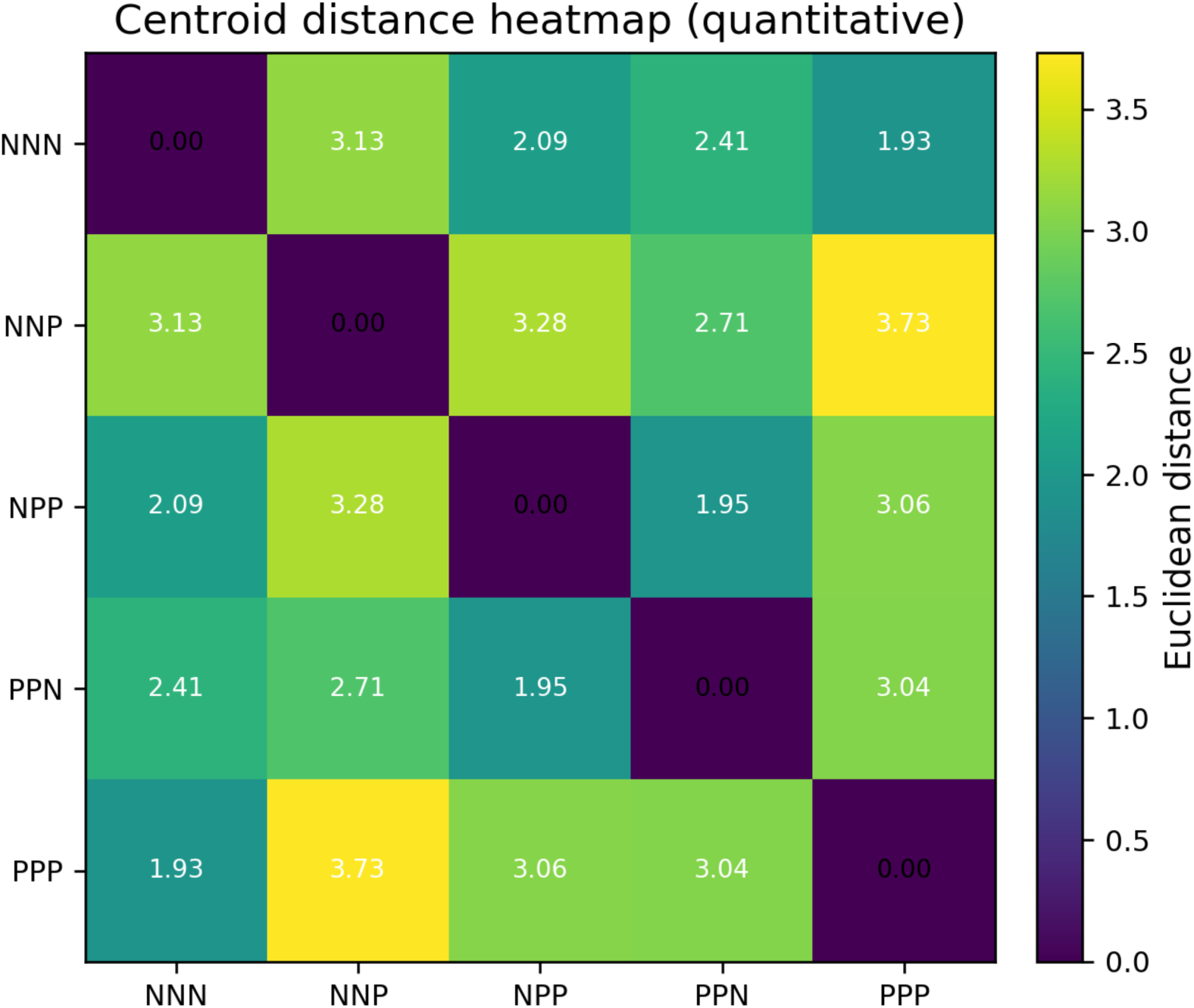
Quantitative centroid distances of quantitative metabolites across classes.

**Supplementary Figure 7:**
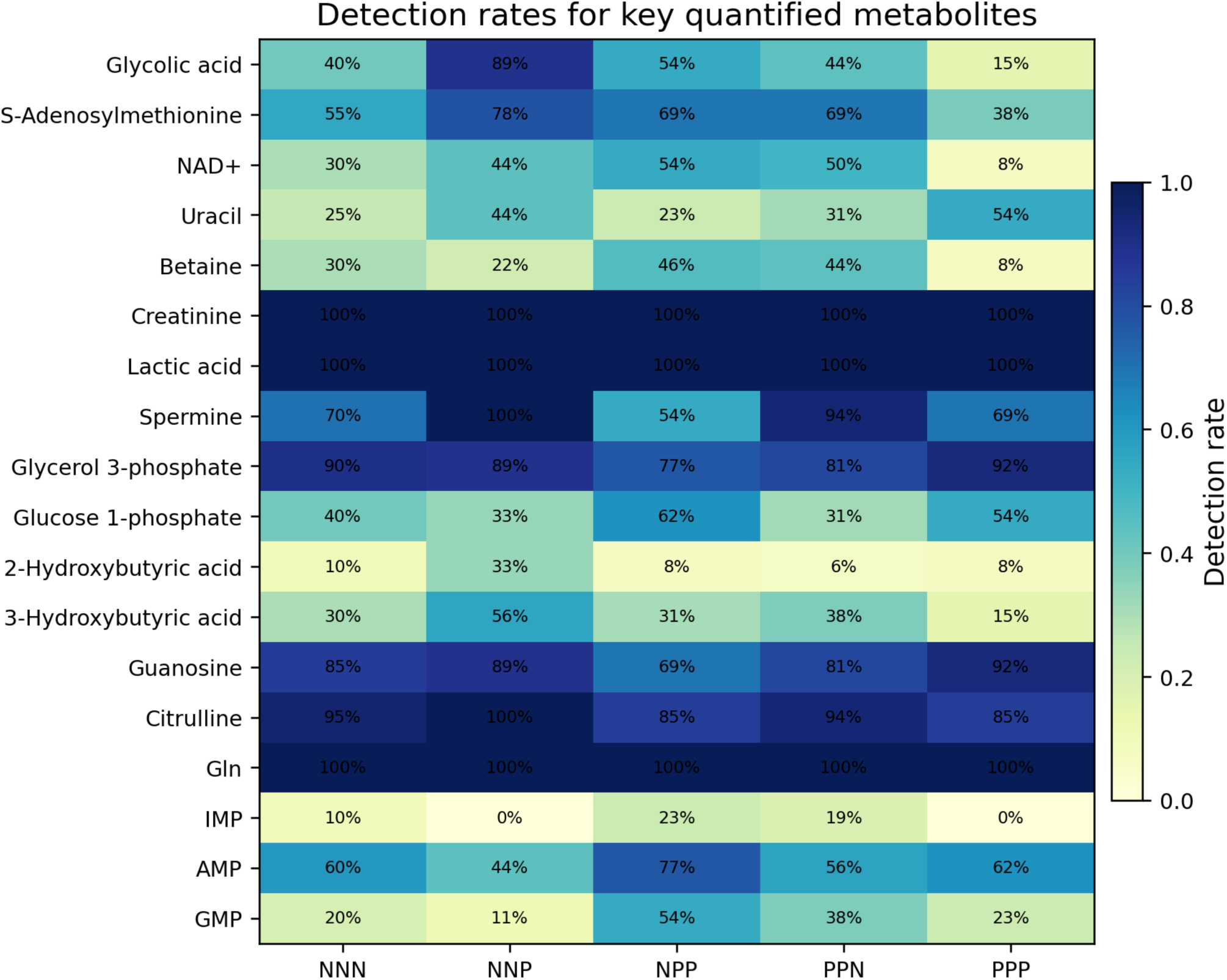
Detection rates for the quantitative metabolites across classes.

